# Metabolomic profiling identifies plasma protein biomarkers of biological aging

**DOI:** 10.64898/2026.09.27.26364110

**Authors:** Xinting Luo, Chong Liu, Yanyu Xie, Yu Zhao, Hanzhu Zhao, Yuanfeng Huang, Shuo Gao, Yijing Wang, Xingxing Jian, Guihu Zhao, Ying Zhang, Jinchen Li, Julian Mutz, Bin Li

## Abstract

Blood metabolomics capture variation in biological aging, but the circulating proteins that underlie or accompany metabolomic aging and its links to disease and mortality remain underexplored. Here, we integrated metabolomics, proteomics and genetics from UK Biobank to characterize the protein architecture of metabolomic aging. Building on our previous mortality-trained score (MetScore1), we developed MetScore2 from 325 NMR biomarkers in 400,594 participants. Higher MetScore2 values marked earlier onset of age-related diseases, and faster score increases predicted higher mortality. Proteome-wide association study identified MetScore2-associated plasma proteins. Their signature was broadly similar in both sexes but showed undulating changes across mid-to-late adulthood, with peaks at 50, 57 and 65 years. By integrating GWAS, Mendelian randomization and colocalization, we identified IL1RN, ARG1, NCAN, F11R, SERPINF2 and NBL1 as putative causal proteins, each with distinct disease associations. Our findings delineate the proteomic landscape of metabolomic aging and prioritize candidate causal proteins for mechanistic and translational follow-up.

## Introduction

People of the same chronological age can differ markedly in disease risk and survival^1,2^. Circulating metabolites provide a systemic readout of this heterogeneity because they reflect genetic background, lifestyle, medication use and tissue metabolism^3,4^. Large biobank studies have shown that plasma metabolite profiles are associated with disease phenotypes, mortality and other lifespan-related traits^5–8^, and metabolomic age models have been linked to morbidity, frailty and survival^9,10^. In our previous work, we developed a 54-biomarker, mortality-trained metabolomic aging score in UK Biobank (hereafter, MetScore1), which was associated with age-related disease and outperformed other aging metrics in predicting short-term mortality risk^9^. However, the circulating protein systems that underlie or accompany metabolomic aging, and their relevance to disease and mortality, remain underexplored.

Plasma proteins offer a route to interpret metabolomic aging because many circulating proteins are secreted, membrane-associated or extracellular molecules with annotated cellular sources and pathway functions^11,12^. Recent plasma-proteomic studies have mapped age-related protein changes across lifespan, organ-aging, frailty and disease cohorts^4,11,13,14^. These studies show that the plasma proteome carries age-related information, but they do not establish which proteins and pathways accompany a mortality-trained metabolomic aging phenotype. Defining this proteomic signature could help connect metabolomic aging to immune signaling, extracellular remodeling and potential circulating therapeutic targets.

Genetic evidence can help distinguish protein associations that are more likely to lie on a disease-relevant pathway from correlations driven by age, lifestyle, medication use or comorbidity. For circulating proteins with protein Quantitative Trait Locus (pQTL) instruments, Mendelian randomization (MR) can test whether genetically predicted protein abundance is associated with a downstream phenotype, and colocalization can assess whether the protein and phenotype associations share the same regional genetic signal^11,12^. Applied to metabolomic aging, this strategy can narrow a broad protein-association map to a smaller set of genetically supported candidates for disease-focused follow-up.

Building on our 54-biomarker MetScore1^9^, we integrated baseline NMR metabolomic profiles, plasma proteomics and genetic data to investigate the protein architecture of metabolomic aging. Initially, we developed MetScore2 as a mortality-trained measure of metabolomic aging in 400,594 UK Biobank participants and evaluated its relationships with age-related disease onset, incident disease risk and longitudinal change. Next, we identified plasma proteins associated with MetScore2 and characterized their pathway enrichment, protein-interaction structure, sex-specific associations and age-related trajectories. Finally, we used Genome-wide Association Study (GWAS), fine-mapping, MR, colocalization and FinnGen phenome-wide MR to prioritize circulating protein candidates and connect them with age-related disease endpoints.

## Results

### MetScore2 represents a mortality-trained metabolomic aging phenotype

To quantify metabolomic aging, we trained MetScore2, a 325-feature NMR biomarker model, to predict all-cause mortality in 400,594 UK Biobank participants (Table 1 and Extended Data Fig. 1). During a median follow-up of 16.68 years, 46,494 deaths occurred. Ten-fold cross-validation generated out-of-fold predictions for all downstream analyses. The strongest contributors were albumin, GlycA, creatinine, glucose, 3-hydroxybutyrate and glutamine (Supplementary Fig. 2a,b).

**Table 1.** Comparison of study design between MetScore1 and MetScore2. MetScore1 refers to the 54-biomarker mortality-trained metabolomic aging score reported in our 2024 study. LASSO, least absolute shrinkage and selection operator.ss

|  | **MetScore1** | **MetScore2** |
| --- | --- | --- |
| UK Biobank sample | 250,341 participants | 400,594 participants |
| NMR feature universe | 325 NMR biomarkers | 325 NMR biomarkers |
| Score construction | 54 biomarkers selected by LASSO Cox | LightGBM with 10-fold cross-validation and out-of-fold predicted MetScore2 |
| WGS sample | 95,372 WGS participants | 398,364 WGS participants |
| Longitudinal repeat-NMR sample | 13,263 participants | 16,974 participants |

### MetScore2 outperforms MetScore1 and chronological age in mortality discrimination

We first assessed how well MetScore2 discriminated all-cause mortality across follow-up time. Time-dependent ROC analyses showed the strongest discrimination for near-term mortality. The out-of-fold AUC reached 0.785 at 1 year and declined gradually to 0.708 at 15 years (Fig. 2a and Supplementary Data 1).

**Figure 1.**
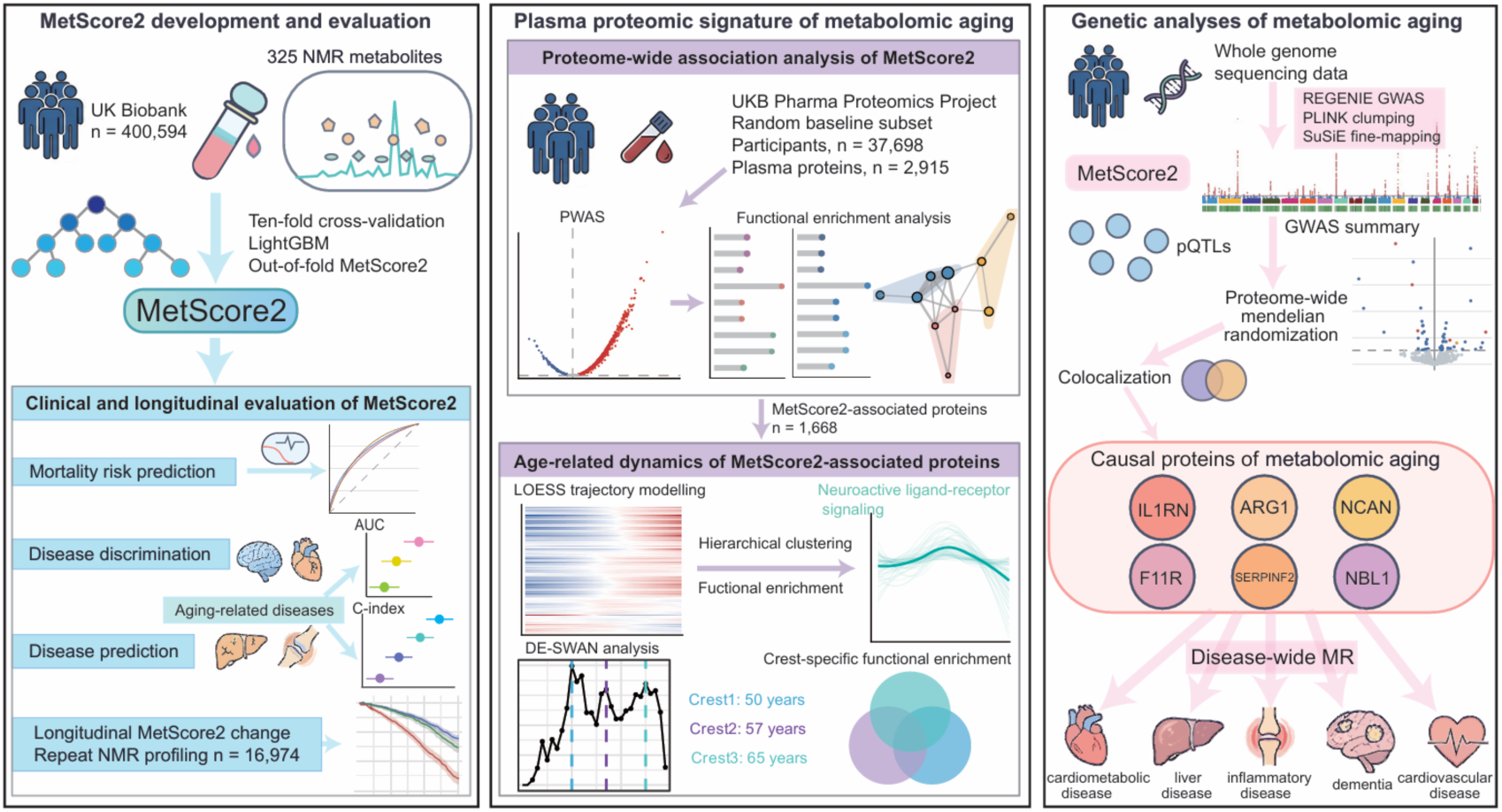
Development, proteomic characterization and genetic analyses of metabolomic aging. **Left**, MetScore2 was developed in 400,594 UK Biobank participants using 325 nuclear magnetic resonance (NMR) metabolites. A mortality-trained LightGBM model was fitted using ten-fold cross-validation and isotonic calibration to generate out-of-fold MetScore2 values. Its clinical performance was evaluated for mortality risk prediction, discrimination of 19 age-related diseases, incremental disease prediction and longitudinal change in a repeat-NMR subset (n = 16,974). **Middle**, PWAS of MetScore2 was conducted in the random baseline subset of the UK Biobank Pharma Proteomics Project (UKB-PPP, n = 37,698; 2,915 plasma proteins). Functional enrichment analysis was performed for MetScore2-associated proteins (n = 1,668). Their age-related dynamics were further characterized using LOESS trajectory modelling, hierarchical clustering, functional enrichment and differential expression sliding-window analysis (DE-SWAN), identifying functional transitions centered at approximately 50, 57 and 65 years. **Right**, whole-genome sequencing (WGS) data were used for MetScore2 GWAS with REGENIE, PLINK clumping and SuSiE fine-mapping. MetScore2 genetic associations were integrated with plasma protein pQTLs through proteome-wide MR and colocalization analyses. Six candidate causal proteins of metabolomic aging were identified: IL1RN, ARG1, NCAN, F11R, SERPINF2 and NBL1. Disease-wide MR was subsequently used to evaluate their associations with cardiometabolic, liver, inflammatory, neurodegenerative and cardiovascular diseases.

**Figure 2.**
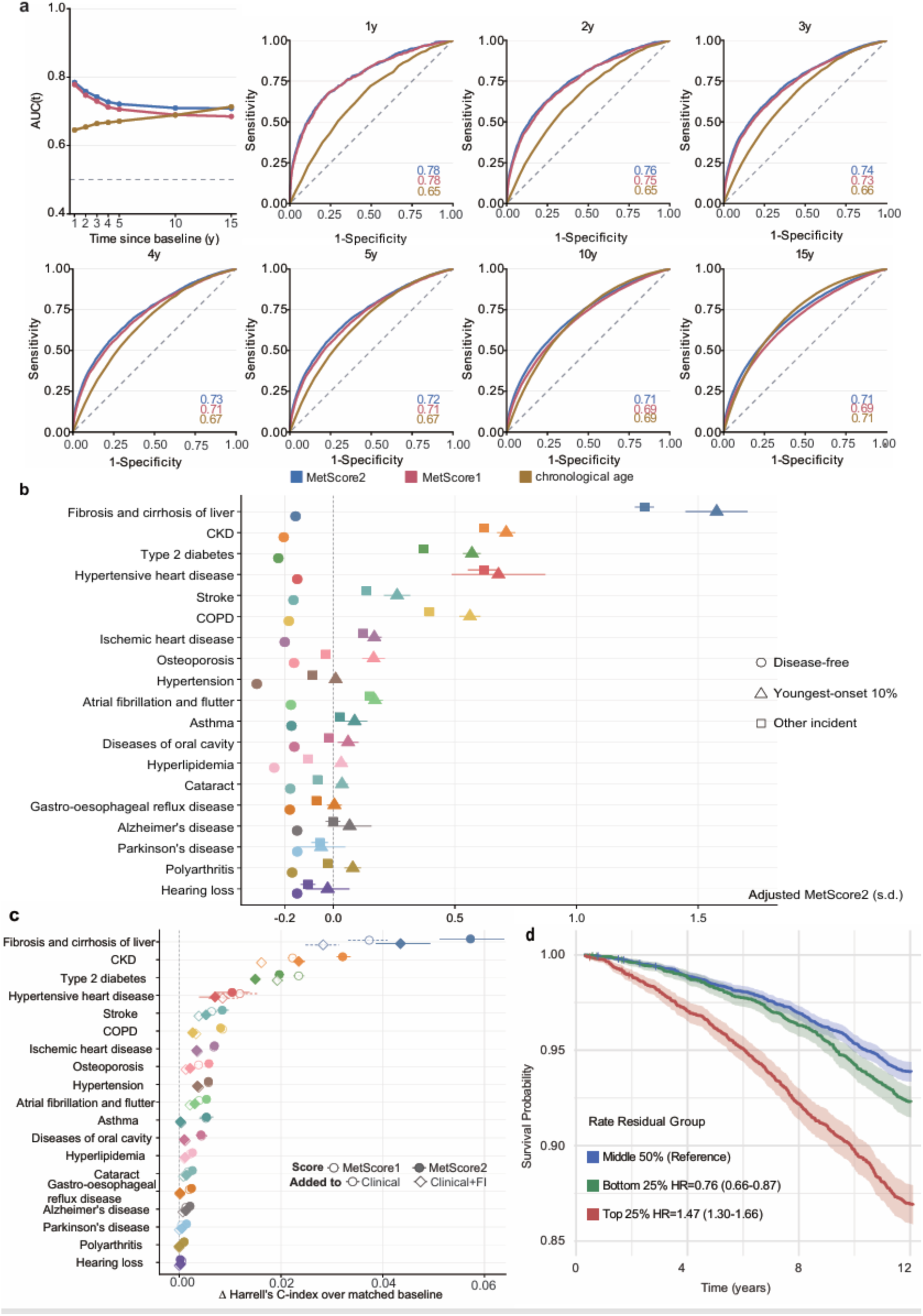
MetScore2 improves mortality discrimination, marks earlier onset of age-related disease and tracks mortality risk over time. **a,** Out-of-fold time-dependent ROC analyses for all-cause mortality at 1, 2, 3, 4, 5, 10 and 15 years after baseline. Blue, MetScore2; red, MetScore1; ochre, chronological age. Dashed diagonal lines indicate chance discrimination. Analyses included 400,594 participants and 46,494 deaths. **b,** Age and sex-adjusted estimated marginal means of MetScore2 for disease-free participants, the youngest decile of age at diagnosis and other incident cases across 19 age-related diseases. Error bars show 95% CI. Participants with prevalent disease were excluded separately for each endpoint; sample sizes are provided in Supplementary Data 2. **c,** Increment in Harrell’s C-index after adding MetScore1 or MetScore2 to the clinical model, with or without the FI. The clinical model included age, sex, BMI, TDI, smoking status, alcohol intake frequency and assessment center. Error bars show 95% bootstrap CI from 200 resamples. **d,** Kaplan-Meier survival estimates by residualized annual MetScore2 change among 16,974 participants with repeat NMR profiling. Lower quartile, middle 50% and upper quartile were classified as slow, medium and fast increase. Shaded bands show 95% CI; crosses indicate censoring. Cox models were adjusted for baseline MetScore2, age at repeat NMR profiling and sex; the middle group was the reference.

Across the same follow-up horizons, MetScore2 showed higher AUCs than MetScore1, including 0.785 versus 0.778 at 1 year and 0.708 versus 0.685 at 15 years. MetScore1 itself outperformed several established aging metrics in our 2024 study^9^. Its advantage over chronological age was largest at early follow-up and became smaller over longer horizons. This time pattern indicates that MetScore2 was most sensitive to deaths occurring close to blood sampling, consistent with a metabolomic measure of current physiological risk.

### Higher MetScore2 marks earlier onset and incidence of age-related diseases

We then examined whether higher MetScore2 values preceded the clinical onset of age-related disease. Across 19 incident diseases, participants diagnosed in the youngest age-at-diagnosis decile had higher baseline MetScore2 values than disease-free participants for 18 endpoints after adjustment for age and sex (Fig. 2b and Supplementary Data 2). The largest elevations were observed for fibrosis and cirrhosis of the liver, chronic kidney disease (CKD), hypertensive heart disease, type 2 diabetes (T2D) and chronic obstructive pulmonary disease (COPD). Disease-specific plots showed weaker separation for several neurological or sensory endpoints, including Parkinson’s disease and hearing loss (Extended Data Fig. 2a). This pattern indicates that MetScore2 captured early disease-related metabolic changes more clearly for hepatic, renal, cardiometabolic and respiratory endpoints.

In prospective Cox models adjusted for clinical covariates and the frailty index (FI), each 1-s.d. higher baseline MetScore2 was associated with incident fibrosis and cirrhosis of the liver (HR 1.67, 95% CI 1.63-1.71), CKD (HR 1.50, 95% CI 1.48-1.51) and T2D (HR 1.32, 95% CI 1.30-1.33; Supplementary Data 2).

We next tested whether MetScore2 improved disease-risk prediction when added to conventional clinical models. Adding MetScore2 increased Harrell’s C-index across all 19 diseases over the clinical baseline model and retained incremental value for 18 diseases after inclusion of the FI (Fig. 2c; disease-specific estimates are shown in Extended Data Fig. 2b). The largest gains appeared for fibrosis and cirrhosis of the liver and CKD, with C-index increases of 0.057 and 0.032 over the clinical model, and 0.043 and 0.023 after adding the FI. These endpoints were supported across three analyses: pre-diagnostic MetScore2 elevation, prospective disease risk and incremental prediction.

After evaluating disease prediction, we examined whether MetScore2 was associated with modifiable clinical, social and lifestyle risk factors. This analysis included socioeconomic status, physical measures, local environment, early-life and reproductive health, diet, psychosocial factors and lifestyle. Higher MetScore2 values were most strongly associated with Townsend deprivation index (TDI), body mass index (BMI), systolic blood pressure and smoking status (Supplementary Fig. 3), all of which are closely linked to chronic disease and mortality.

### Faster MetScore2 increase is associated with higher mortality risk

We next examined whether change in MetScore2 over time was related to mortality. Among 16,974 participants with repeat NMR profiling, the median interval between measurements was 4.5 years. We calculated annualized MetScore2 change between the two visits and adjusted this change for baseline MetScore2, age at repeat profiling and sex. This adjustment separated score increase during follow-up from the participant’s initial MetScore2.

We then divided participants into three groups using the residualized annual change in MetScore2: the fastest 25%, the middle 50% and the slowest 25%. Participants in the fastest-increase group had a higher subsequent mortality hazard than the middle group (HR 1.47, 95% CI 1.30-1.66; Fig. 2d). Participants in the slowest-increase group had a lower mortality hazard (HR 0.76, 95% CI 0.66-0.87).

### MetScore2 is associated with immune and tissue-remodeling proteins

To identify plasma proteins associated with MetScore2, we performed a protein-wide association study (PWAS) of 2,915 plasma proteins in 37,698 UKB-PPP participants. After adjustment for demographic, clinical and genetic covariates, 1,668 proteins reached the Bonferroni threshold (Fig. 3a and Supplementary Data 3). Associations were mainly positive, with 1,476 proteins increasing and 192 decreasing in abundance with higher MetScore2 values. The largest positive associations were observed for GDF15 (β = 0.452, 95% CI 0.442-0.462), TNFRSF1A (β = 0.372, 95% CI 0.363-0.382), WFDC2 (β = 0.357, 95% CI 0.348-0.367), IGFBP4 (β = 0.346, 95% CI 0.337-0.356), SHISA5 (β = 0.343, 95% CI 0.334-0.352) and VSIG4 (β = 0.340, 95% CI 0.331-0.349). The strongest inverse associations included PON3 (β = -0.219), FGFBP1 (β = -0.192), APOC1 (β = -0.182), APOM (β = -0.181) and PLA2G7 (β = -0.179).

**Figure 3.**
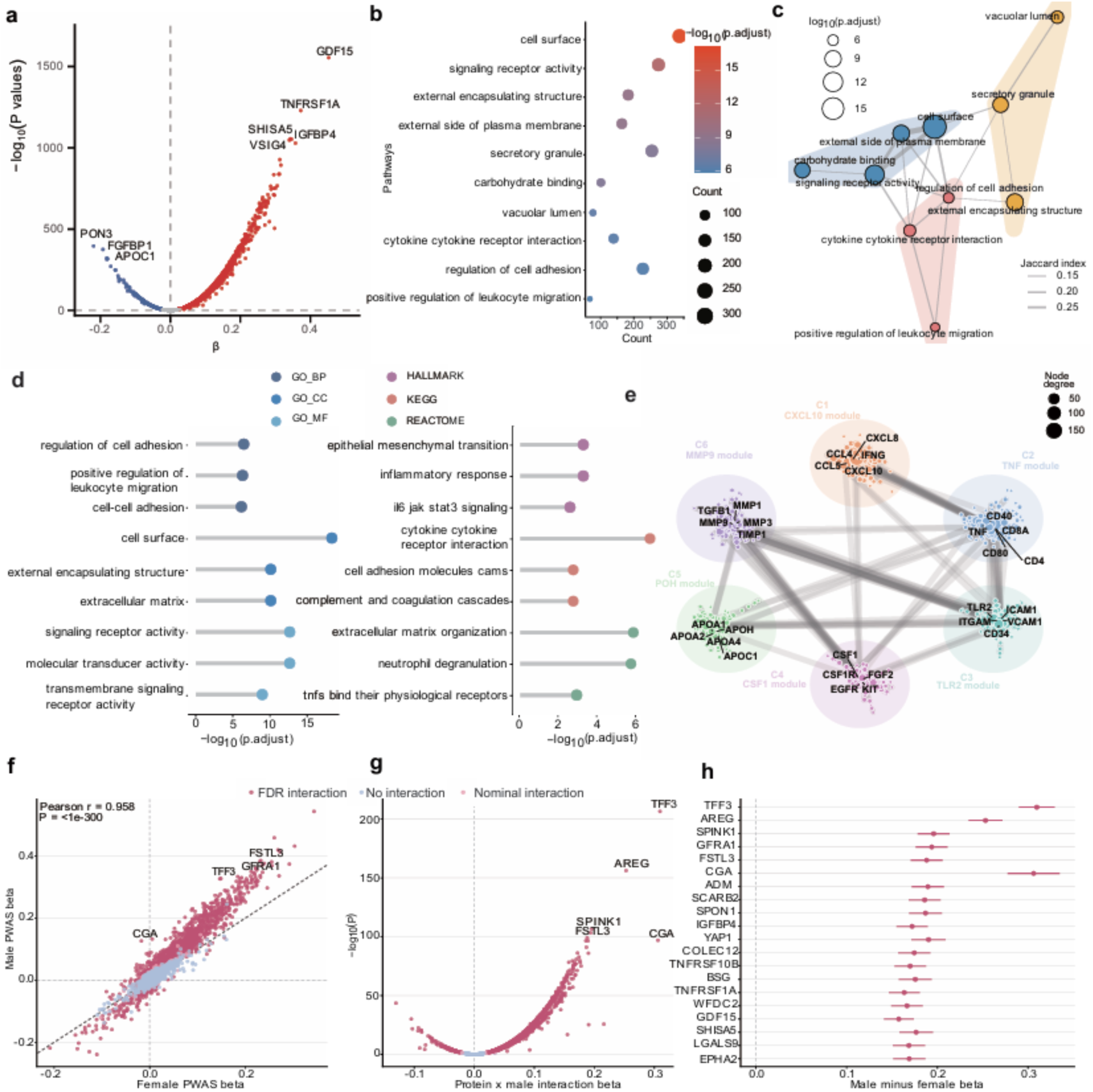
Plasma proteomic profiles of MetScore2. **a,** Proteome-wide associations between standardized protein abundance and out-of-fold MetScore2 in 37,698 UKB-PPP participants. Each point represents one of 2,915 Olink proteins. Beta values indicate adjusted differences in MetScore2 per 1-s.d. higher protein abundance; dashed lines mark beta = 0 and the Bonferroni threshold (P = 1.72 × 10^-5^). **b,** Representative Gene Ontology terms enriched among 1,668 Bonferroni-significant proteins. Point size and colour denote protein count and adjusted P value. **c,** Enrichment map. Nodes represent terms, node size denotes adjusted significance, edges denote gene-set Jaccard overlap and colours denote Louvain communities. **d,** Selected enriched terms across Gene Ontology, Hallmark, KEGG and Reactome. **e,** Six highest-ranked maximal clique centrality communities in the STRING interaction network. Nodes are proteins sized by degree; edges are STRING interactions; labelled nodes are top-ranked proteins. **f,** Sex-stratified PWAS effect estimates. Each point represents one protein; the diagonal indicates equal estimates and dashed lines mark zero. **g,** Protein-by-sex interaction estimates; positive values indicate stronger associations in males. **h,** Top 20 protein-by-sex interactions.

We next examined which biological pathways were over-represented among the MetScore2-associated proteins. The strongest Gene Ontology biological-process terms included regulation of cell adhesion (227 proteins, adjusted P = 9.84 × 10^-7^), positive regulation of leukocyte migration (68 proteins, adjusted P = 1.39 × 10^-6^), cell-cell adhesion (269 proteins, adjusted P = 1.94 × 10^-6^), chemotaxis (159 proteins, adjusted P = 1.94 × 10^-6^) and leukocyte migration (147 proteins, adjusted P = 1.94 × 10^-6^) (Fig. 3b,d and Supplementary Data 3). Cellular-component enrichment pointed to cell surface (337 proteins, adjusted P = 8.93 × 10^-18^), extracellular matrix (ECM, 183 proteins, adjusted P = 4.09 × 10^-10^), external side of plasma membrane (164 proteins, adjusted P = 6.50 × 10^-10^) and secretory granule (254 proteins, adjusted P = 5.58 × 10^-9^) (Fig. 3b,c). KEGG and Reactome analyses gave the same direction of evidence, with enrichment for cytokine-cytokine receptor interaction, ECM organization, neutrophil degranulation, complement/coagulation cascades and TNF-receptor binding. Together, these results show that a higher MetScore2 was accompanied by higher activity of immune-cell recruitment, cell-surface signaling, secretion and extracellular-matrix turnover in the circulating plasma protein.

A list of 1,668 proteins is too broad to interpret directly, so we used STRING to examine whether the associated proteins formed connected groups. Of the Bonferroni-significant proteins, 1,341 were connected by 7,346 STRING edges (Fig. 3e and Supplementary Data 4). The highest-ranked hubs included CXCL10, CXCL8, CCL5, CCL4, IFNG, IL6, CXCL9, IL1B, IL10 and TNF. The main modules contained chemokine/cytokine proteins (CXCL10, CXCL8, CCL5, IFNG and IL6), T-cell and TNF-related proteins (TNF, CD4, CD8A, CD40 and CD80), myeloid and adhesion proteins (TLR2, ICAM1, ITGAM and VCAM1), growth-factor signaling proteins (CSF1, KIT, EGFR and CSF1R), matrix-remodeling proteins (MMP9, MMP3, TGFB1 and TIMP1) and lipid/coagulation-related proteins (APOH, APOA1, APOA2, APOC1 and APOM). Thus, network analysis reduced the PWAS results to connected systems involving inflammatory signaling, immune-cell adhesion, matrix remodeling and lipid/coagulation biology.

Because males and females differ in average mortality risk and prevalence of many cardiometabolic diseases, we repeated the PWAS stratified by sex to test whether the main MetScore2-protein associations were present in both groups. The protein-MetScore2 associations were highly similar between sexes (Pearson r = 0.958) and 2,456 of 2,915 proteins showed the same direction of association in males and females (Fig. 3f). The same leading proteins also remained strongly positive in both groups. For example, GDF15 was associated with MetScore2 in females (β = 0.343) and males (β = 0.543), and TNFRSF1A was also associated with MetScore2 in females (β = 0.303) and males (β = 0.431).

Protein-by-sex interaction analysis identified 1,465 proteins at 5% FDR and 953 proteins at the Bonferroni threshold (Fig. 3g,h and Supplementary Data 3). The largest male-predominant differences included TFF3 (females β = 0.149; males β = 0.327; interaction P = 2.77 × 10^-207^), AREG (females β = 0.146; males β = 0.326; interaction P = 5.43 × 10^-157^), SPINK1 (females β = 0.184; males β = 0.333; interaction P = 2.21 × 10^-107^), GFRA1, FSTL3, ADM, SCARB2, WFDC2 and GDF15. Several inverse associations were also stronger in males, including PLA2G7, APOM, TTR and PON1. These findings suggest that the broad immune and tissue-remodeling protein signature of MetScore2 is shared by males and females, whereas the magnitude of individual protein associations is often sex dependent.

Points and horizontal lines denote beta values and 95% CI. All P values were two-sided. Multiple testing used the Benjamini-Hochberg procedure. Additional PWAS, enrichment and sex-interaction results are provided in Supplementary Data 3; network data are provided in Supplementary Data 4.

### Undulating transitions in the MetScore2-associated proteome in middle and later life

To characterize age-related changes in the MetScore2-associated proteome, we estimated LOESS-smoothed protein trajectories from 40 to 70 years for the 1,668 Bonferroni-significant proteins. Hierarchical clustering identified six trajectory groups: two increasing clusters, one decreasing cluster, two midlife peak-or-trough clusters and one cluster that increased from early to midlife and then declined at older ages (Fig. 4a-d and Supplementary Data 5). Non-monotonic patterns were observed for 301 proteins (18.0%) and were enriched for oxidative phosphorylation, peroxisome and neuroactive ligand-receptor signaling.

**Figure 4.**
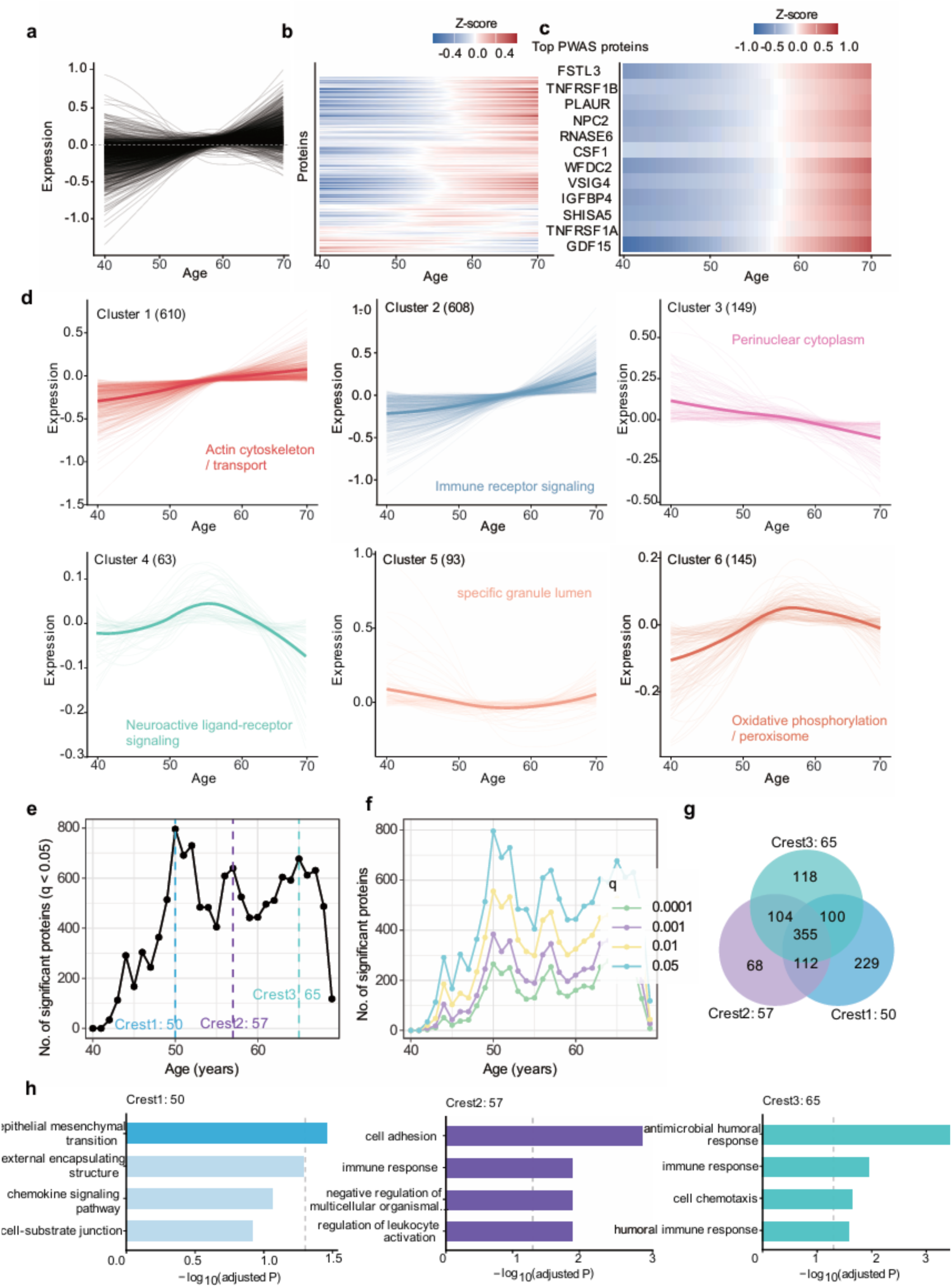
Undulating age-related changes in the MetScore2-associated plasma proteome. **a,** LOESS-smoothed age-related expression patterns for the 1,668 Bonferroni-significant MetScore2-associated proteins from ages 40 to 70 years. Fitted values are shown on the standardized protein-abundance scale. **b,** Heatmap of normalized fitted protein trajectories across chronological age. **c,** Age-related fitted trajectories for selected top MetScore2-associated proteins from the PWAS. **d,** Six trajectory clusters identified from normalized LOESS-fitted age profiles, with representative enriched pathways annotated for each cluster. **e,** Number of proteins significantly different between the younger and older halves of each sliding age window in DE-SWAN analysis at q < 0.05. Peaks mark the three leading transition crests centered at approximately 50, 57 and 65 years. **f,** Significant protein counts across FDR thresholds from q < 0.0001 to q < 0.05. **g,** Overlap of proteins contributing to the three leading age-window crests. **h,** Representative enriched terms for crest-specific proteins, grouped by crest. Enrichment analyses used the tested MetScore2-associated proteins as the background set. Multiple testing in DE-SWAN and enrichment analyses was controlled using the Benjamini-Hochberg procedure. Full trajectory assignments, crest-level protein lists and enrichment results are provided in Supplementary Data 5. Source data are provided with this paper.

We next applied DE-SWAN to identify and quantify nonlinear changes in the MetScore2-associated proteome across adulthood. We found three crests around ages 50, 57 and 65, involving 796, 639 and 677 significant proteins, respectively (Fig. 4e and Supplementary Data 5). Most significant proteins were higher in the older half of each window (769 of 796, 624 of 639 and 614 of 677), and the same crest pattern was retained across FDR thresholds from q < 0.0001 to q < 0.05 (Fig. 4f).

To further examine the biological processes represented by each crest, we performed separate enrichment analyses for crest-specific proteins. The three crests shared 355 proteins but differed in their enriched terms: epithelial-mesenchymal transition, extracellular-matrix and adhesion terms around age 50; signaling receptor activity, cell adhesion and adaptive immune pathways around age 57; and cytokine, chemokine, antimicrobial humoral response and extracellular-matrix terms around age 65 (Fig. 4g,h and Supplementary Data 5).

### Proteome-wide MR analyses identify IL1RN, ARG1, NCAN, F11R, SERPINF2 and NBL1 as potential causal proteins of MetScore2

After identifying plasma proteins associated with MetScore2, we performed genetic analyses to identify circulating proteins that may contribute to metabolomic aging. We first examined the genetic architecture of MetScore2 outside the UKB-PPP sample to avoid overlap with the proteomic data used in downstream MR.

MetScore2 GWAS of 6,868,881 WGS variants identified 78 significant loci and 185 independent lead variants, with a median analytical sample size of 355,628 participants per variant (Fig. 5a and Supplementary Data 6). Although the lambdaGC was 1.453, the lambda1000 was 1.001 and the LD score regression intercept was 1.077, indicating that the association signal was largely attributable to polygenicity (Fig. 5b). Fine-mapping further resolved these loci: 69 of 78 contained at least one credible set, and 14 had a top variant with PIP ≥ 0.95 (Supplementary Fig. 5 and Supplementary Data 6).

**Figure 5.**
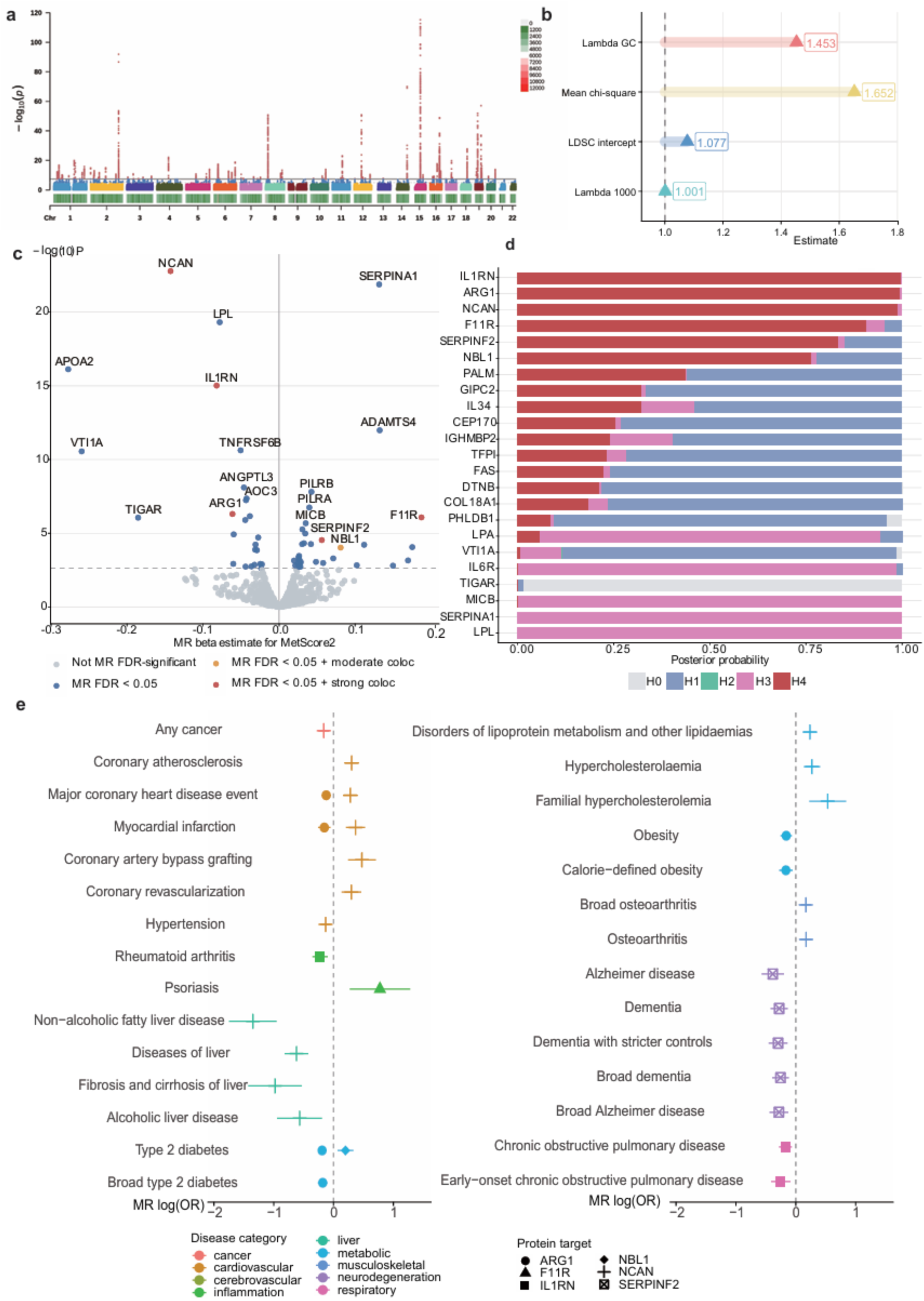
Genetic analyses prioritize circulating proteins linked to MetScore2. **a,** Manhattan plot of the GWAS of standardized out-of-fold MetScore2 after excluding UKB-PPP participants. Each point represents one WGS variant; the horizontal line indicates genome-wide significance (P = 5 × 10^-8^). **b,** Genomic inflation and polygenicity diagnostics. LambdaGC, genomic-control inflation factor; lambda1000, sample-size-standardized genomic-control inflation factor; LDSC, linkage-disequilibrium score regression. **c,** Proteome-wide MR estimates for genetically predicted circulating protein abundance and MetScore2. Each point represents one tested protein. The x axis shows the MR beta estimate for MetScore2 and the y axis shows - log10(P). The dashed line indicates the 5% FDR threshold. Colors denote MR and colocalization status. **d,** Colocalization posterior probabilities for MR-significant proteins with sufficient harmonized regional variants. H0-H4 denote no association, protein-only association, MetScore2-only association, distinct signals and shared signal, respectively. Strong and moderate colocalization support were defined as PP.H4 ≥ 0.80 and PP.H4 ≥ 0.50. **e,** FinnGen disease-wide MR results for the six prioritized proteins. Points and horizontal lines indicate MR log(OR) estimates and 95% CI. Colors denote disease category and symbols denote protein target. Only associations passing global FDR correction across 480 target-disease pairs are shown. GWAS and fine-mapping results are in Supplementary Data 6, MR and colocalization results in Supplementary Data 7, and FinnGen results in Supplementary Data 8.

After performing the MetScore2 GWAS, we used MR to identify plasma proteins with potential causal links with MetScore2. Using cis-pQTL instruments for 1,108 proteins, this analysis identified 57 proteins associated with MetScore2 after FDR correction (Fig. 5c, Extended Data Fig. 3 and Supplementary Data 7). The strongest inverse MR associations included NCAN (β = - 0.143, P = 1.66 × 10^-23^), IL1RN (β = -0.082, P = 9.66 × 10^-16^) and ARG1 (β = -0.061, P = 4.94 × 10^-7^). The strongest positive associations among proteins later supported by colocalization included F11R (β = 0.187, P = 8.42 × 10^-7^), NBL1 (β = 0.081, P = 9.41 × 10^-5^) and SERPINF2 (β = 0.056, P = 2.91 × 10^-5^).

We then used colocalization to identify colocalized MR signals with MetScore2. Among the 57 MR-associated proteins, 23 had sufficient harmonized regional variants for colocalization. Six proteins showed moderate or strong evidence of colocalization: IL1RN (PP.H4 = 0.998), ARG1 (PP.H4 = 0.994), NCAN (PP.H4 = 0.987), F11R (PP.H4 = 0.908), SERPINF2 (PP.H4 = 0.834) and NBL1 (PP.H4 = 0.763) (Fig. 5d, Extended Data Fig. 4 and Supplementary Data 7). These analyses prioritized IL1RN, ARG1, NCAN, F11R, SERPINF2 and NBL1 as candidate protein drivers of metabolomic aging.

### Six prioritized proteins show distinct disease associations

We then tested whether the six prioritized proteins also showed evidence of causal links with age-related disease endpoints. Using FinnGen GWAS summary statistics, disease-wide MR across 480 target-disease pairs identified 32 associations after global FDR correction (Fig. 5e, Supplementary Fig. 6 and Supplementary Data 8). Genetically higher ARG1 was associated with lower risks of T2D (OR = 0.83, 95% CI 0.77-0.88), obesity (OR = 0.85, 95% CI 0.78-0.94), major coronary heart disease (OR = 0.88, 95% CI 0.82-0.96) and myocardial infarction (OR = 0.86, 95% CI 0.78-0.95). Genetically higher IL1RN was associated with lower risks of rheumatoid arthritis (OR = 0.80, 95% CI 0.71-0.91), COPD (OR = 0.84, 95% CI 0.76-0.93) and early-onset COPD (OR = 0.78, 95% CI 0.66-0.92).

F11R and NBL1 showed positive disease associations matching their positive associations with MetScore2. Genetically higher F11R was associated with higher risk of psoriasis (OR = 2.17, 95% CI 1.31-3.59), and genetically higher NBL1 was associated with higher risk of T2D (OR = 1.22, 95% CI 1.07-1.39). NCAN showed the broadest disease profile. Genetically higher NCAN was associated with lower risks of non-alcoholic fatty liver disease (OR = 0.26, 95% CI 0.17-0.39), liver disease (OR = 0.54, 95% CI 0.44-0.66) and liver fibrosis/cirrhosis (OR = 0.38, 95% CI 0.24-0.59), but with higher risks of coronary atherosclerosis (OR = 1.35, 95% CI 1.20-1.53), myocardial infarction (OR = 1.44, 95% CI 1.23-1.69) and lipoprotein disorders (OR = 1.27, 95% CI 1.13-1.42). Genetically higher SERPINF2 was associated with lower risks of Alzheimer’s disease (OR = 0.68, 95% CI 0.57-0.82) and dementia (OR = 0.76, 95% CI 0.66-0.87). These disease-wide results separated the six prioritized proteins into distinct disease categories. ARG1 and IL1RN pointed mainly to lower cardiometabolic and inflammatory risk, F11R and NBL1 to higher psoriasis or T2D risk, NCAN to opposing liver and coronary/lipid associations and SERPINF2 to lower dementia-related risk.

## Discussion

We developed MetScore2, a mortality-trained metabolomic aging score based on 325 NMR biomarkers, in 400,594 UK Biobank participants. Participants who developed age-related diseases at younger ages had higher baseline MetScore2 values, and adding MetScore2 improved risk prediction of 19 endpoints. Faster increases in MetScore2 over repeat profiling predicted higher subsequent mortality. Integrating PWAS with genetics prioritized IL1RN, ARG1, NCAN, F11R, SERPINF2 and NBL1 as candidate causal proteins of metabolomic aging, providing targets for experimental validation and mechanistic studies.

MetScore1 showed that a blood-based, mortality-trained aging score was associated with mortality, age-related disease risk and longitudinal aging rate^9^, but the circulating proteins that underlie or accompany this signal remained unclear. Here, we updated the score in a larger UK Biobank sample and integrated it with plasma proteomics and genetic analyses, including PWAS, MR and colocalization.

Recent large-scale metabolomic studies have shown that plasma metabolites can predict common diseases and map disease-related genetic signals. For example, Buergel et al. showed that NMR metabolomic profiles predicted most common diseases and added predictive information for T2D, dementia and heart failure^15^. Barrett et al. further supported the value of circulating metabolites for disease-risk prediction across populations^16^. You et al. then linked plasma metabolites to disease risk, metabolomic risk scores and colocalized genetic associations at population scale^7^. We extend these studies by using metabolomic aging as the phenotype for plasma protein discovery and genetic target prioritization.

Blood metabolites are useful for quantifying biological aging because they reflect lipid transport, amino-acid metabolism, glucose handling, inflammation, kidney function and nutrition. In this study, we defined metabolomic aging as a plasma metabolite pattern linked to mortality and age-related disease. People with a higher MetScore2 had lower albumin^17^ and higher GlycA^18^, creatinine, glucose, 3-hydroxybutyrate and glutamine, and this pattern reflects inflammation, kidney stress, abnormal glucose-ketone metabolism and poorer systemic reserve. Higher GlycA reflects systemic inflammation^19^, and lower albumin has been linked to poorer survival in older adults^17^. Disease and repeat-measurement analyses showed that this state can appear before clinical diagnosis and can worsen over time. This interpretation is consistent with recent metabolomic age and metabolome-phenome studies linking blood metabolites to frailty, mortality, lifespan-related traits and future disease risk^7,10,15^.

The plasma metabolome is well suited for clinical follow-up because it can be measured from blood and reflects current physiological state. MetScore1 used this feature to identify high-risk metabolomic aging before age-related disease diagnosis^9^. In the present study, MetScore2 preserved this clinical signal in a larger cohort. Repeated profiling added a dynamic readout: faster increases in MetScore2 were followed by higher mortality. These findings supported its use as the phenotype for plasma protein biomarker discovery and causal protein prioritization.

These findings also help clarify what MetScore2 reflects. It is best interpreted as a mortality-trained measure of current physiological risk, which helps explain a pattern seen in both MetScore1 and MetScore2: discrimination was strongest for near-term mortality and became weaker over longer follow-up^9^. The leading contributors to MetScore2, including albumin, GlycA, creatinine and glucose, point to inflammation, organ dysfunction and metabolic stress as major components of this signal. At the same time, MetScore2 was elevated before diagnosis across 18 of 19 age-related diseases and its rate of change predicted subsequent mortality, suggesting that the score also contains information that precedes clinical diagnosis. The modest gain in mortality discrimination over MetScore1 should also be viewed in this context. AUC and C-index often change little when a new marker is added to an already informative model, even when the marker is associated with the outcome^20–22^. Similar modest gains have been reported for blood-biomarker biological-age models in UK Biobank^23^. In addition, all-cause mortality is influenced by clinical history, lifestyle, socioeconomic factors and incident disease events, which are only partly measured by baseline NMR metabolites^24,25^.

Our PWAS linked metabolomic aging to inflammatory, vascular and tissue-remodeling pathways. At the protein level, the leading network hubs included CXCL10, CXCL8, CCL5, CCL4, IFNG, IL6, IL1B, IL10 and TNF, placing cytokine and chemokine signaling at the center of the MetScore2 protein map^26–28^. Cell-adhesion and leukocyte-migration pathways connected this immune signal to endothelial interaction and immune-cell entry into tissues^29–31^. Extracellular-matrix pathways linked metabolomic aging to tissue repair and scarring^32–34^. Complement and coagulation pathways further suggested disturbed clot formation and clot resolution^35,36^. This biology gives a clearer meaning to high metabolomic aging, marking a metabolic state tied to inflammation, vascular injury, tissue scarring and impaired clot resolution.

Recent plasma-proteomic studies have shown that blood proteins capture different aging phenotypes. Lehallier et al. reported non-linear plasma-protein waves across the lifespan^4^. Proteomic aging clocks used plasma proteins to estimate biological age and predict mortality, multimorbidity and age-related disease risk^37^. Organ-aging models linked plasma proteins to organ-specific aging and mortality^38^. Brain-aging work linked plasma proteins to imaging-derived brain age and prioritized BCAN as a candidate biomarker^39^. Frailty proteomics connected clinical vulnerability to plasma proteins, MR-prioritized candidates and age-related protein waves^34^. Our study uses metabolomic aging as the phenotype for protein discovery, linking a mortality-related metabolic state to plasma protein biomarkers, pathways and candidate causal proteins.

Our study identifies IL1RN, ARG1, NCAN, F11R, SERPINF2 and NBL1 as candidate causal proteins of metabolomic aging. Among these proteins, IL1RN showed the strongest colocalized protective association with metabolomic aging. IL1RN encodes the interleukin-1 receptor antagonist, and loss of IL1RN causes severe systemic autoinflammation^40^. Human genetic studies also showed that IL1RN-raising alleles reduce IL-6 and C-reactive protein^41^. In our disease-wide analysis, genetically higher IL1RN was associated with lower rheumatoid arthritis and COPD risk. Together, these observations prioritize IL1RN for experimental studies of inflammation in metabolomic aging. F11R and ARG1 may connect metabolomic aging to endothelial dysfunction. Genetically predicted higher F11R was associated with higher MetScore2. F11R (JAM-A) is expressed by platelets and inflamed endothelial cells, where it contributes to platelet adhesion, leukocyte trafficking and endothelial interactions^42,43^. This interpretation is consistent with the enrichment of cell-adhesion and leukocyte-migration pathways in our PWAS. Arginase limits L-arginine availability for nitric oxide synthesis, and increased arginase activity has been linked to endothelial dysfunction and vascular disease^44–46^. In our analysis, higher genetically predicted ARG1 was associated with lower MetScore2 and lower cardiometabolic disease risk. NBL1, a secreted BMP antagonist previously linked to diabetic kidney disease^47,48^, was another genetically prioritized candidate and its contribution to metabolomic aging requires experimental clarification. Together with IL1RN, these findings implicate inflammatory, endothelial, arginine-metabolic and tissue-repair pathways for disease-specific investigation.

Notably, not all of the genetically prioritized proteins showed the same pattern across diseases. NCAN and SERPINF2 are two examples. Genetically predicted higher NCAN was associated with lower MetScore2 and lower risks of liver disease and cirrhosis in our phenome-wide analyses. The NCAN-CILP2-TM6SF2 region is known to influence hepatic steatosis, inflammation, fibrosis and circulating lipid traits^49–51^. In particular, TM6SF2 variation can increase liver fat while reducing the secretion of triglyceride-rich lipoproteins^50–52^. The effects of this locus may therefore differ between liver disease and cardiovascular risk.

By contrast, genetically predicted higher SERPINF2 was associated with higher MetScore2. Alpha-2-antiplasmin, encoded by SERPINF2, inhibits fibrinolysis and stabilizes fibrin clots^35,53^. Experimental studies have also linked alpha-2-antiplasmin to microvascular thrombosis and poorer outcomes after cerebral thromboembolism^36^. However, higher predicted SERPINF2 was associated with lower risks of Alzheimer’s disease and dementia in our analysis. These results show that NCAN and SERPINF2 should be tested separately in each disease setting before they are considered as therapeutic targets.

Another key finding of our study is that proteins associated with metabolomic aging changed unevenly across adulthood. We observed three peaks at approximately 50, 57 and 65 years. Similar non-linear patterns have been reported in several studies, including general plasma proteome waves across the lifespan^4^, brain-aging protein waves around ages 57, 70 and 78^39^, and frailty-associated protein crests around ages 50 and 63^34^. The differing peak ages across studies likely reflect the distinct aging phenotypes that each capture. In our data, the age-50 peak was enriched for extracellular-matrix and cell-adhesion pathways, the age-57 peak for receptor signaling and adaptive immunity, and the age-65 peak for cytokine, chemokine and antimicrobial-response pathways. These results suggest that the protein profile of metabolomic aging shifts from tissue remodeling in midlife toward stronger immune and inflammatory signals later in life.

There are certain limitations to our study. First, plasma protein concentrations do not directly measure protein expression or pathway activity in specific tissues. Plasma assays are more likely to capture secreted, extracellular and immune-related proteins, and may miss intracellular metabolism, mitochondrial function and tissue-resident repair. Second, the proteomic associations and age-related protein trajectories were estimated from cross-sectional proteomic data. Thus, the LOESS and DE-SWAN analyses identify between-participant age-associated patterns and peaks in proteomic change, but cannot determine whether the same patterns occur longitudinally within individuals or establish discrete biological transition ages. Independent cohorts with repeated metabolomic and proteomic measurements are needed to test the reproducibility and temporal ordering of these findings. Third, MR and colocalization help prioritize proteins but do not show that changing protein levels will change MetScore2 or reduce disease risk. Although we used cis-pQTL instruments and excluded UKB-PPP participants from the MetScore2 GWAS to minimize sample overlap, horizontal pleiotropy through neighbouring genes cannot be fully excluded, and colocalization support varied across targets. IL1RN, F11R, ARG1, NBL1, NCAN and SERPINF2 should therefore be tested experimentally, with attention to tissue-specific effects, direction of effect and safety. Fourth, analyses were restricted to UK Biobank participants of White British genetic ancestry, who were mainly middle-aged or older and healthier than the general population. Both MetScore2 and the pQTL instruments used here therefore require validation in cohorts of other ancestries and age ranges before wider application.

In conclusion, our study shows that MetScore2 captures a circulating metabolic profile associated with mortality and earlier onset of age-related disease. Participants with more rapid increases in MetScore2 had a higher subsequent mortality risk. By integrating NMR metabolomics, plasma proteomics and genetic analyses, we linked this profile to several protein pathways and identified six circulating proteins (IL1RN, F11R, ARG1, NBL1, NCAN and SERPINF2) as candidate causal proteins of metabolomic aging. These findings support the use of metabolomic aging to prioritize protein targets for age-related disease. Future studies combining repeated metabolomic and proteomic profiling in diverse cohorts with tissue-specific perturbation experiments should test whether these proteins influence MetScore2 progression and age-related disease risk.

## Methods

### Study population and omics data

We analyzed participants from the UK Biobank, a prospective cohort of approximately 500,000 adults aged 40-69 years recruited between 2006 and 2010. The UK Biobank has ethical approval from the North West Multi-centre Research Ethics Committee, and all participants provided written informed consent. This study was conducted under UK Biobank application ID 103082. Analyses were restricted to participants of White British genetic ancestry (UKB field ID: 22006) with baseline plasma NMR metabolomic data, a valid baseline assessment date (UKB field ID: 53), positive follow-up time and at least 80% non-missing processed metabolite measures. Mortality follow-up was defined from baseline assessment to death (UKB field ID: 40000), loss to follow-up (UKB field ID: 191) or administrative censoring on 20 November 2025, whichever occurred first.

### NMR metabolomic profiling and preprocessing

Baseline NMR metabolomic data were obtained from EDTA plasma samples assayed by the UK Biobank Nightingale Health Phase 3 release. Technical and batch effects were corrected using the ukbnmr R package (version 3). Following correction, 107 non-derived metabolomic biomarkers were retained, and 218 additional features were derived, comprising 61 composite measures, 22 ratios and 135 percentages, yielding 325 features for analysis. Absolute-concentration measures and ratios were natural-log transformed as log(x + m/2), where m is the feature-specific observed minimum; percentage measures were not transformed. All features were then standardized to z scores. Values recorded below or above the assay quantification limits were retained. No imputation was undertaken during initial metabolomic preprocessing because feature-level missingness was below 3%.

### UKB-PPP plasma proteomic data and preprocessing

Baseline plasma proteomic data were obtained from the UKB-PPP, which measured circulating proteins in plasma using the Olink Explore proximity extension assay platform.

Protein abundance was represented by normalized protein expression (NPX), a relative log2-scale measure.

As UKB-PPP includes both a random baseline sample and consortium-selected or disease-enriched participants, all primary proteomic analyses were restricted to the random baseline subset overlapping the primary MetScore2 cohort, and consortium-selected participants were excluded. Proteins with more than 20% missing values were excluded (8 of 2,923 proteins), and participants with more than 80% missing values across retained proteins were removed (53 participants). Remaining missing protein values were imputed using k-nearest-neighbour imputation (k = 10), after which each protein was standardized to z scores. The final primary proteomic dataset included 37,698 participants and 2,915 proteins.

### WGS data and genetic analysis sample

WGS data were used for genetic analyses of MetScore2^54^. Among participants in the primary metabolomic analysis cohort, 398,364 participants had available WGS data and were included in genome-wide analyses after genetic quality control. To minimize sample overlap with UKB-PPP protein pQTL data in downstream MR analyses, participants with baseline UKB-PPP proteomic measurements were excluded from the MetScore2 GWAS used as the outcome dataset for proteome-wide MR and colocalization.

### Construction of MetScore2

MetScore2 was constructed as a metabolite-only mortality score. Age, sex, BMI, TDI, smoking, alcohol intake and other clinical variables were not used as model inputs. Participants were divided into ten folds stratified by mortality status, sex (UKB field ID: 31) and baseline age (UKB field ID: 21003) group. In each outer fold, the model was trained in the other nine folds and then applied to the held-out fold, so each participant received a MetScore2 value from a model that had not been trained on that participant.

Within each training set, candidate models used the top 30, 54, 100, 200 or 325 NMR metabolite features. Feature ranking was obtained from a LightGBM classifier fitted in the training data, and the feature number was selected using an internal 20% validation split^55^. The final fold-specific model used LightGBM with isotonic calibration from five-fold cross-validation. Selected features could differ across outer folds because feature ranking and feature-number selection were repeated within each training set. Missing metabolite values were imputed using medians from the corresponding outer training fold before model fitting or prediction. The predicted mortality probability from each held-out fold was standardized within that fold to generate MetScore2.

### Comparison of MetScore2 with MetScore1 and chronological age for mortality prediction

Mortality prediction was evaluated using held-out MetScore2 values. MetScore1 was recalculated in the present cohort using the 54-metabolite coefficient set reported in our previous study and was standardized before comparison with MetScore2 and chronological age^9^. MetScore2, MetScore1 and chronological age were compared for all-cause mortality prediction at 1, 2, 3, 4, 5, 10 and 15 years after baseline. Time-dependent AUCs were estimated with cumulative dynamic ROC analysis. Cox proportional-hazards models were used to estimate mortality associations per 1-s.d. higher score.

### Baseline MetScore2 differences across youngest-onset, other-onset and disease-free groups

We constructed 19 aging-related disease endpoints from linked UK Biobank health records. Disease-specific UK Biobank first-occurrence date fields and ICD-10 code groups are listed in the Supplementary Tables. For each endpoint, participants with disease before or on the baseline assessment date were excluded. Follow-up time was calculated from baseline to the first disease record, death, loss to follow-up or administrative censoring. Among incident cases, participants in the youngest 10% of age at diagnosis were classified as youngest-onset cases. Remaining incident cases were classified as other-onset cases, and participants without the disease during follow-up were classified as disease-free.

Age and sex-adjusted baseline MetScore2 levels were compared across disease-free participants, youngest-onset cases and other-onset cases using linear models. Pairwise contrasts were calculated from the fitted model, and P values were adjusted with the Benjamini-Hochberg method.

### Incremental disease-risk prediction beyond conventional clinical models

Disease-risk prediction was evaluated using Cox models trained in nine folds and tested in the held-out fold. The base clinical model included age (UKB field ID: 21003), sex (UKB field ID: 31), body mass index (BMI; UKB field ID: 21001), Townsend deprivation index (TDI; UKB field ID: 22189), smoking status (UKB field ID: 20116), alcohol intake frequency (UKB field ID: 1558) and assessment center (UKB field ID: 54). We compared the base clinical model with models adding MetScore1, MetScore2, the FI, the FI plus MetScore1 or the FI plus MetScore2. The FI definition is provided in the Supplementary Methods. Harrell’s C-index was calculated from held-out linear predictors^56^. CI and differences in C-index were estimated with 200 bootstrap resamples. Endpoints with fewer than 200 eligible participants or fewer than 20 incident cases were not modelled.

### Longitudinal change in MetScore2

Participants with repeat NMR metabolomic measurements were scored using the trained MetScore2 models. Annual change was calculated as the difference between repeat and baseline MetScore2 divided by the time interval between visits. To reduce dependence on the baseline score, annual change was regressed on baseline MetScore2, baseline age and sex. The residual from this model was used as the adjusted rate of MetScore2 change. Participants in the lowest quartile, middle two quartiles and highest quartile were labelled as slow, medium and fast changers. Landmark mortality follow-up began at the repeat NMR visit. Cox models compared slow and fast changers with medium changers, adjusting for age at repeat assessment and sex. We checked proportional-hazards assumptions using scaled Schoenfeld residuals for all Cox models.

### Associations of modifiable risk factors with MetScore2

We tested 71 baseline modifiable risk factors for association with MetScore2, corresponding to 253 overall factor-level contrasts. Continuous variables were z-scored. Ordinal and categorical variables were modelled as factor-level contrasts against predefined reference levels listed in the Supplementary Tables. The primary model adjusted for age, sex, assessment center and the first 20 genetic principal components (PCs). P values were adjusted with the Benjamini-Hochberg method within each subgroup analysis. These analyses were interpreted as baseline associations, not intervention effects.

### PWAS of MetScore2

PWAS used baseline UKB-PPP Olink Explore NPX data from the random baseline subset overlapping with the MetScore2 cohort^57^. Consortium-selected or disease-enriched UKB-PPP participants were excluded from the primary analysis. Proteins with more than 20% missing values were removed, participants with more than 80% missing retained proteins were excluded, remaining missing values were imputed by k-nearest neighbors imputation with k = 10 and each protein was z-scored^58^. After these preprocessing steps, the primary PWAS included 37,698 participants and 2,915 proteins.

For each protein, we fitted a linear model with MetScore2 as the outcome and z-scored protein level as the exposure. Model 1 adjusted for age, sex and the first 20 genetic PCs (UKB field ID: 22009). Model 2 additionally adjusted for available behavioral and socioeconomic covariates, including BMI, TDI, smoking and alcohol intake. The reported beta represents the difference in MetScore2 s.d. units per 1-s.d. higher protein. Bonferroni correction across tested proteins defined the primary significance threshold, and Benjamini-Hochberg FDR was also reported.

### Pathway enrichment of MetScore2-associated proteins

Functional enrichment used Bonferroni-significant MetScore2-associated proteins as the query set and all tested PWAS proteins as the background. Olink assay names were mapped to gene symbols, and multi-gene assay names were split into separate symbols. Gene Ontology enrichment was performed with clusterProfiler and org.Hs.eg.db^59^. Hallmark, KEGG and Reactome enrichment used MSigDB GMT files^60–63^. Terms with adjusted P ≤ 0.05 were retained.

### STRING protein-protein interaction (PPI) analysis

Bonferroni-significant MetScore2-associated proteins were submitted to STRING for Homo sapiens PPI analysis^64^. We used the full STRING network, a high-confidence combined interaction score threshold of 0.700 and no additional interactors. STRING node and edge exports were merged with the PWAS results, and the retained interactions were analysed as an undirected weighted graph in R using igraph^65^. Hub proteins were ranked by maximal clique centrality, and protein modules were assigned by Louvain community detection.

### Age-related protein trajectory analysis

Age trajectories were assessed for Bonferroni-significant MetScore2-associated proteins in the random baseline UKB-PPP subset. Each protein was z-scored before trajectory analysis. Cross-sectional trajectories from age 40 to 70 years were estimated using LOESS. Proteins were clustered by hierarchical clustering with Ward linkage using the LOESS trajectories.

### DE-SWAN age-window analysis

DE-SWAN was used to identify ages at which local protein differences were concentrated^66^. Age was rounded to integer years. For each age center, participants in the younger and older halves of a 6-year window were compared, corresponding to [c - 3, c) versus [c, c + 3). Models were adjusted for sex, the first 20 genetic PCs, TDI, smoking status, alcohol intake frequency and BMI. Crests were defined from local maxima in the number of FDR-significant proteins, requiring a minimum 4-year separation between selected crests. Crest-specific enrichment used tested DE-SWAN proteins as the background.

### Sex-stratified PWAS sensitivity analysis

Sex-stratified PWAS was performed as a sensitivity analysis. The primary stratified model was fitted separately in males and females and included z-scored protein level, age, the first 20 genetic PCs and the model-2 covariates available in each stratum. Sex was omitted from the stratified models. A separate interaction model included protein, sex and a protein-by-sex interaction term, with the interaction beta interpreted as the male-minus-female difference in the protein-MetScore2 association. Protein values were z-scored within each analysis sample.

### GWAS of MetScore2

GWAS used MetScore2 as a quantitative phenotype. Participants with baseline Olink proteomic data were excluded before GWAS input generation to reduce sample overlap with UKB-PPP pQTL instruments, and finally the GWAS had a median variant-level sample size of 355,628 participants. REGENIE was used for two-step association testing^67^. Covariates included age, sex, age squared, age-by-sex interaction, age-squared-by-sex interaction, the first 20 genetic PCs and assessment center. Analyses used autosomal variants passing genetic quality control which required minor allele frequency ≥ 0.01, Hardy-Weinberg equilibrium P > 1 × 10^-6^, genotype missingness ≤ 0.03 and minor allele count ≥ 100 in REGENIE step 2. Genome-wide significance was set at P < 5 × 10^-8^.

### Post-GWAS fine-mapping

Post-GWAS analyses used the no-proteomics MetScore2 GWAS summary statistics. LDSC was applied to HapMap3-overlapping variants to estimate the intercept, attenuation ratio and SNP heritability^68^. Independent lead variants were reported using PLINK clumping with P1 = 5 × 10^-8^, P2 = 1 × 10^-5^, r^2^ < 0.1 and a 1-Mb window^69^. Fine-mapping used SuSiE RSS within merged 1-Mb windows around genome-wide significant variants^70^. LD matrices were calculated from the no-proteomics UK Biobank genotype subset. SuSiE was run with up to ten effects and 95% credible sets.

### Proteome-wide MR

Proteome-wide MR tested whether genetically predicted plasma protein abundance was associated with MetScore2. Cis-pQTL instruments were extracted from UKB-PPP European pQTL summary statistics within 1 Mb of the encoding gene using GENCODE v49 annotations^71^. The primary analysis used independent genome-wide significant cis-pQTL instruments with F statistic > 10 after LD clumping at r2 < 0.001 within a 10-Mb window; relaxed pQTL thresholds were used in sensitivity analyses. Ambiguous palindromic variants were removed. One-instrument analyses used the Wald ratio; analyses with at least two instruments used inverse-variance weighting (IVW); analyses with at least three instruments also used MR-Egger^72^, weighted median^73^ and weighted mode^74^. P values were adjusted within instrument set and MR method using Benjamini-Hochberg FDR.

### Colocalization analysis

Colocalization was performed for proteins significant in the primary MR tier, defined by genome-wide significant cis-pQTL instruments, Wald-ratio MR and FDR < 0.05. For each protein, pQTL and MetScore2 GWAS variants were extracted from the same cis region, matched by GRCh38 chromosome and position, harmonized by allele, and analysed using coloc.abf for quantitative traits. PP.H4 ≥ 0.8 was treated as strong evidence for a shared causal variant, and PP.H4 ≥ 0.5 as moderate evidence.

### Disease Phe-MR of prioritized protein targets

Prioritized protein targets were carried forward to disease Phe-MR when they had MR evidence for MetScore2 and colocalization support. We tested whether genetically predicted levels of these proteins were causally associated with curated FinnGen R10 aging-related disease endpoints^75^. The endpoint panel was capped at 80 endpoints with at least 1,000 cases when available. Target cis-pQTL instruments were extracted from each FinnGen endpoint, harmonized to the protein-increasing allele, and analysed using Wald ratio or fixed-effect IVW according to the number of instruments. Estimates were reported as odds ratios per genetically predicted 1-unit higher protein abundance. Benjamini-Hochberg FDR was calculated across all target-endpoint tests.

## Data Availability

The individual-level data used in this study are available from UK Biobank under controlled access. This study was conducted under UK Biobank application 103082. Access to UK Biobank phenotype, metabolomic, WGS and UKB-PPP proteomic data can be requested through the UK Biobank Access Management System (https://www.ukbiobank.ac.uk/register-apply/). Individual-level UK Biobank data cannot be redistributed by the authors. Summary results supporting the main figures and supplementary analyses are provided in the Supplementary Tables and Source Data files. Public annotation resources used in downstream analyses included STRING, MSigDB, Gene Ontology, GENCODE. FinnGen R10 GWAS summary statistics used for disease Phe-MR are available through FinnGen after registration at https://www.finngen.fi/en/access_results, with release-specific documentation available at https://finngen.gitbook.io/documentation/r10.

## Code Availability

Analysis scripts will be made available upon publication at https://github.com/Xinting-Luo/Metabolomic-Aging. Analyses were performed primarily in R versions 4.2.2-4.5.3 and Python version 3.10.20. Key software included LightGBM v4.6.0, REGENIE v4.1.2, PLINK v2.0.0-a.7.5LM, LDSC v1.0.1, susieR v0.12.35, coloc v5.2.3, TwoSampleMR v0.7.3, clusterProfiler v4.18.4 and STRING v12.0.

## Acknowledgements

This research has been conducted using the UK Biobank Resource under Application Number 103082. We thank the UK Biobank participants and staff for making this research possible. We thank the High-Performance Computing Center of Central South University for providing the computing platform. We also thank the Center for Computational Biology and Bioinformatics, Furong Laboratory, Central South University, the Bioinformatics Center, Xiangya Hospital, Central South University, and the Shen Jiyuan (Shanghai) Health Technology Co., Ltd., for their support.

## Author contributions

X.L. designed the analytical strategy, curated the data, performed all analyses, generated the figures and tables, interpreted the results and wrote the original draft. J.M. provided methodological guidance and critically revised the manuscript. Y. Zhang and B.L. critically revised the manuscript. J.L. and B.L. provided supervision and intellectual guidance. C.L., Y.X., Y. Zhao, H.Z., Y.H., S.G., Y.W., X.J. and G.Z. contributed to discussion of the results and reviewed the manuscript. All authors read and approved the final manuscript.

