## Extended Data Fig1, Extended Data Fig2, Extended Data Fig3, Extended Data Fig4 for "Metabolomic profiling identifies plasma protein biomarkers of biological aging"

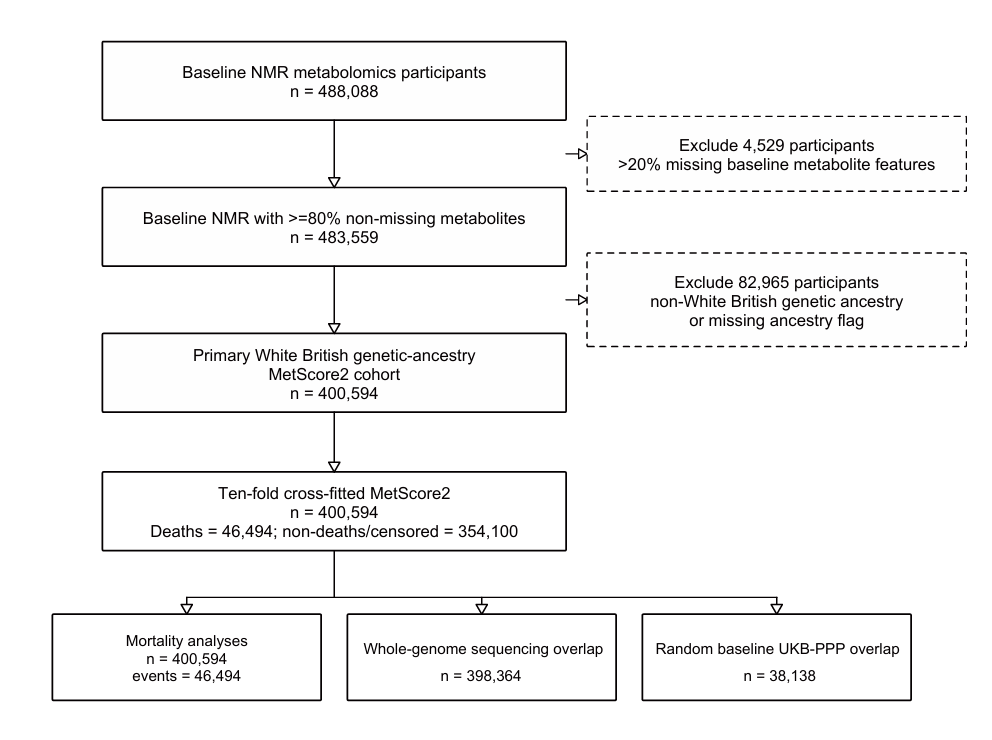
 **Extended Data Fig. 1 | Participant selection and analytical subsets.**

Flow diagram showing the derivation of the UK Biobank NMR metabolomics cohort and the nested analytical subsets used throughout the study. Participants were excluded for missing metabolomic predictors or key covariates, failed metabolomics quality control, ancestry filtering where required, and outcome-specific missingness. The final MetScore2 cohort comprised 400,594 participants, including 46,494 deaths during a median follow-up of 16.68 years. The diagram also indicates the overlap with whole-genome sequencing (WGS) data, the random baseline UKB-PPP proteomics subset and the disease-specific analysis sets. Counts refer to unique participants unless otherwise stated.

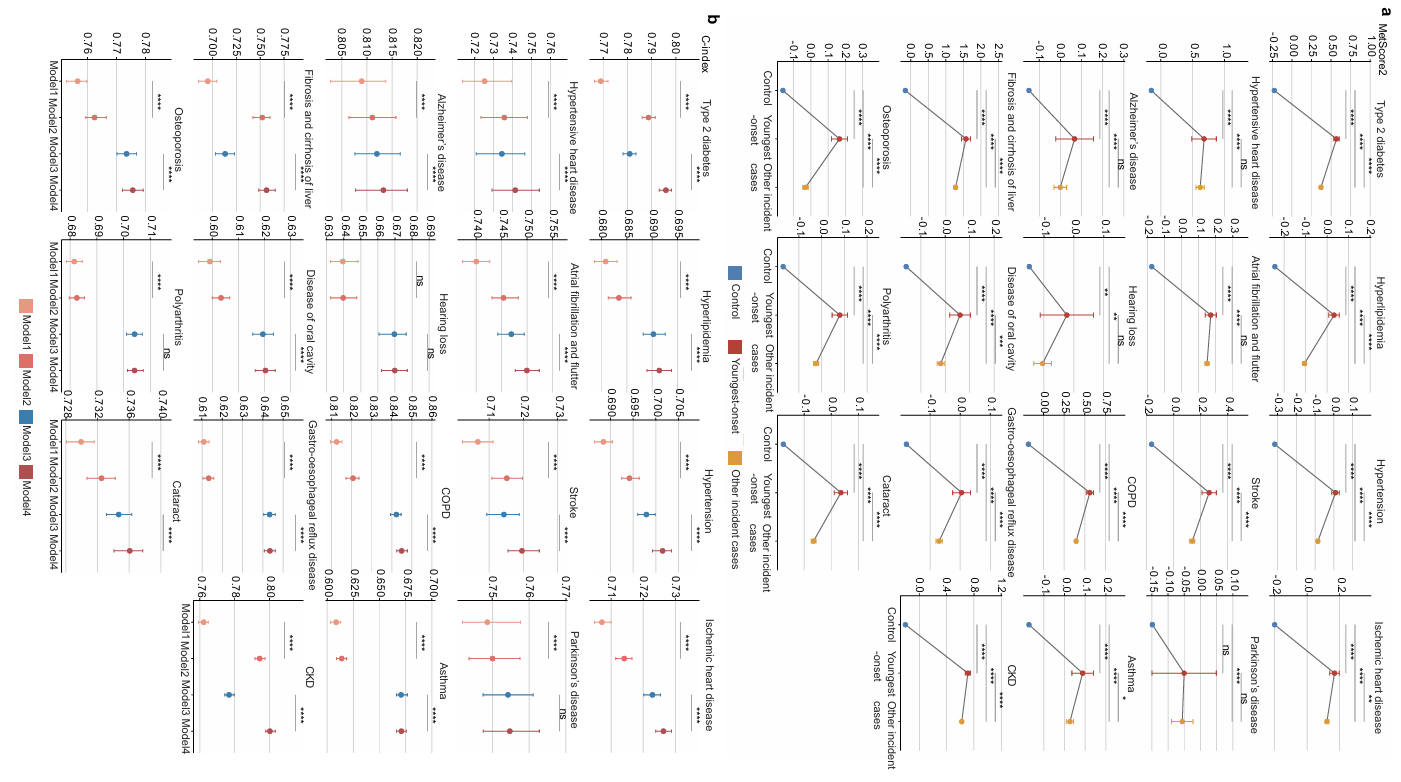

**Extended Data Fig. 2 | MetScore2 marks early disease onset and improves disease-risk prediction.**

**a,** Age- and sex-adjusted MetScore2 levels by disease outcome. **b,** Predictive performance for nested disease-risk models, reported as Harrell's C-index and change in C-index. Points indicate estimates and intervals indicate 95% confidence intervals. Outcome-specific exclusions, case definitions, sample sizes and full model estimates are provided in Supplementary Data 2.

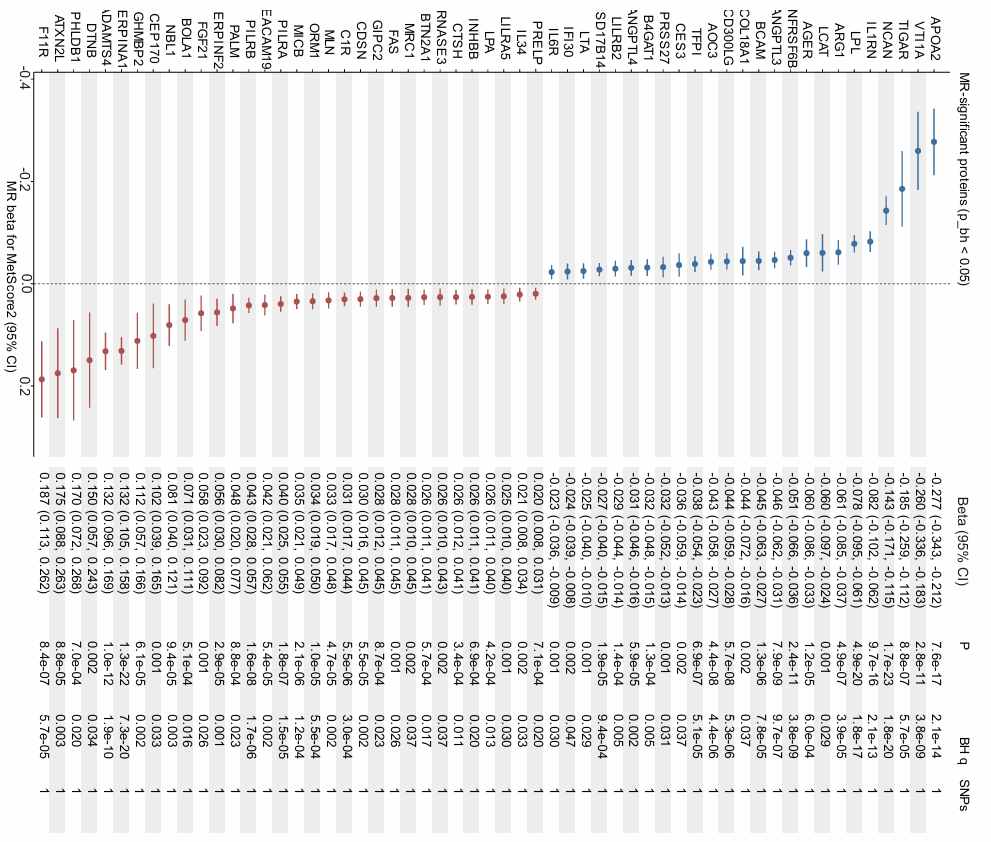
**Extended Data Fig. 3 | Proteome-wide Mendelian randomization signals for MetScore2.**

Forest plot of proteins with evidence of a cis-pQTL-instrumented effect on MetScore2 in proteome-wide Mendelian randomization. Each row shows the estimated effect of genetically predicted protein abundance on MetScore2 with 95% confidence intervals. Proteins are grouped by direction and strength of evidence where shown. Estimates are reported on the MetScore2 scale, and statistical evidence is based on the MR model described in the Methods with false-discovery-rate control across tested proteins. The full set of MR estimates, instrument counts and sensitivity analyses is provided in Supplementary Data 7.

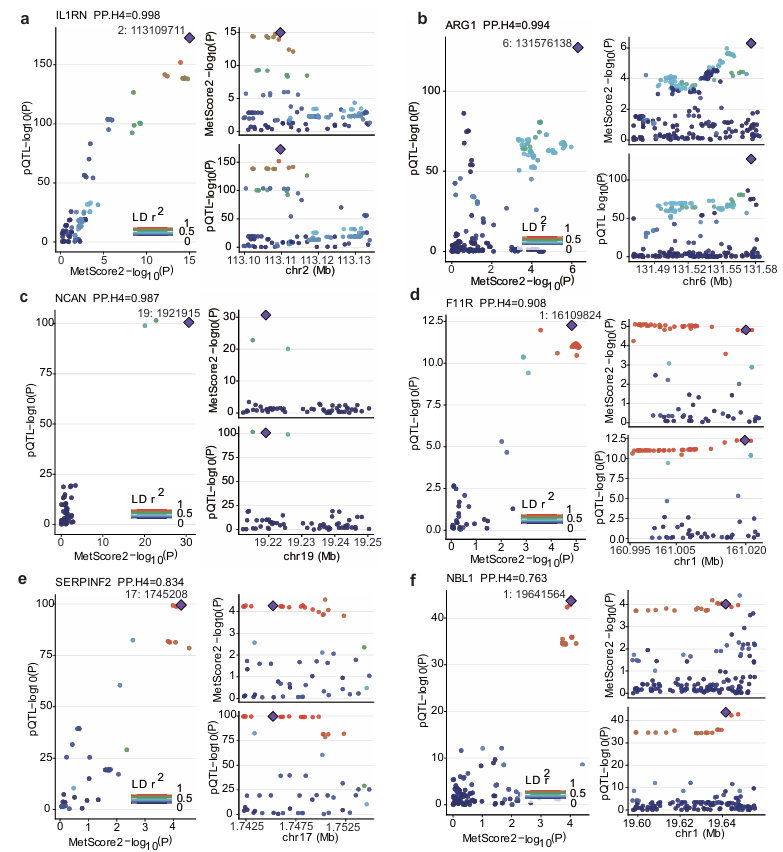
**Extended Data Fig. 4 | Colocalization support for prioritized MetScore2 protein targets.**

Regional colocalization plots for six prioritized protein targets. **a-f,** Locus-level comparison of protein quantitative trait locus and MetScore2 association evidence for IL1RN, ARG1, NCAN, F11R, SERPINF2 and NBL1. Points show variants in the regional window, coloured by linkage disequilibrium to the lead signal where shown. Tracks display the corresponding pQTL and MetScore2 association patterns, and panel labels report posterior support for a shared causal signal. Posterior probabilities for colocalization were PP.H4 = 0.998 for IL1RN, 0.994 for ARG1, 0.987 for NCAN, 0.908 for F11R, 0.834 for SERPINF2 and 0.763 for NBL1. Full MR and colocalization results are provided in Supplementary Data 7.
