## Supplementary Information for "Metabolomic profiling identifies plasma protein biomarkers of biological aging"

**Supplementary Methods**

***Study population and analytical samples***

In addition to the inclusion criteria described in the main Methods, participants were excluded if age or sex was missing, if the recorded death date preceded baseline assessment, or if follow-up time was non-positive. The primary metabolomic cohort was used as the common baseline population for MetScore2 construction, mortality benchmarking, disease analyses and repeat-visit scoring. Analysis-specific missing-data filters were applied before each downstream analysis.

***NMR metabolomic preprocessing***

The processed NMR feature matrix used for MetScore2 was generated before model construction. After technical and batch-effect correction with ukbnmr (version 3), 107 non-derived metabolic biomarkers were retained. We generated a further 218 derived variables, comprising 61 composite measures, 22 ratios and 135 percentage measures, resulting in 325 analysis features. Absolute-concentration measures and ratios were transformed as log(x + m/2), where m was the feature-specific observed minimum; percentage measures were not log-transformed. Features were subsequently standardized to z scores. Values recorded below or above assay quantification limits were retained, and no imputation was applied during this initial preprocessing stage because feature-level missingness was below 3%.

A feature-level data dictionary, including the source variables, derivation formula, transformation and feature class for all 325 measures, is provided in Supplementary Data 1.

***Cross-fitted MetScore2 construction***

All feature ranking, feature-number selection, imputation and calibration steps were performed within the training portion of each outer fold. The held-out fold was used only for prediction.

LightGBM models used a binary objective^2^, 1,000 estimators, learning rate 0.02, 63 leaves, subsampling fraction 0.85, column-sampling fraction 0.90, minimum child samples of 80, L1 regularization of 0.05, L2 regularization of 0.05 and balanced class weights. The random seed was fixed at 20260814 for model training and fold-level procedures.

***Disease endpoint analyses***

Disease-specific first-occurrence fields and ICD-10 code groups are listed in the Supplementary Data. For each endpoint, the first available disease record was used as the diagnosis date. Age at diagnosis was calculated from baseline age and the interval between baseline assessment and the first disease record.

For baseline MetScore2 comparisons, the disease-free group included participants without that endpoint during follow-up after removal of prevalent cases. For disease-risk prediction, endpoint-specific analysis sets were restricted to participants with complete data for the outcome, MetScore2 fold assignment, MetScore2, MetScore1, frailty index and clinical covariates. Cross-fold prediction used the same outer-fold assignments as MetScore2 construction: for each fold, the Cox model was fitted in the other nine folds and evaluated in the held-out fold.

***Frailty index***

The frailty index was calculated from baseline deficit variables listed in the Supplementary Data. For each participant, available deficit components were summed and divided by the number of non-missing components. Frailty index values were set to missing when ten or more deficit components were unavailable or when no deficit component was available. Frailty categories were defined as non-frail for frailty index values <=0.08, pre-frail for values >0.08 and <0.25, and frail for values >=0.25.

***Longitudinal MetScore2 change***

For repeat-visit analyses, the repeat assessment date was used to define the interval between baseline and repeat metabolomic measurement. Participants were excluded from longitudinal analyses if repeat MetScore2 was missing, repeat assessment date was missing or the interval between visits was non-positive. Annualized MetScore2 change was calculated after applying the same processed metabolomic feature space used for baseline MetScore2 scoring.

***Modifiable risk-factor definitions***

We tested 71 baseline modifiable risk factors, corresponding to 253 overall factor-level contrasts. Variable definitions, UK Biobank field IDs, coding rules and reference levels are provided in the Supplementary Data. Variable definitions, UK Biobank field IDs, coding rules and reference levels for modifiable risk factors are provided in the Supplementary Data. Reference levels were chosen before model fitting and were kept fixed across subgroup analyses. For ordinal variables, categories were treated as reported unless a binary exposure definition had been specified in the variable definition.

The healthy-diet indicator was derived from fresh fruit intake (field ID 1309), raw vegetable intake (field ID 1299), cooked vegetable intake (field ID 1289), oily fish intake (field ID 1329), non-oily fish intake (field ID 1339), processed meat intake (field ID 1349), beef intake (field ID 1369), lamb or mutton intake (field ID 1379), and pork intake (field ID 1389).

***UKB-PPP proteomic preprocessing***

Primary proteomic analyses were restricted to the random baseline UKB-PPP subset overlapping the primary MetScore2 cohort. The random-baseline eligibility flag was used where available; otherwise, consortium-selected or disease-enriched participants were excluded using UK Biobank field 30903. Released Olink Explore normalized protein expression values were used following UKB-PPP/Olink quality control, with no additional outlier removal.

After restricting to participants with available MetScore2 values and required model covariates, protein assays with more than 20% missing values were excluded (8 of 2,923 proteins). Participants with more than 80% missing values across the retained assays were then excluded (53 participants). Remaining missing protein values were imputed using k-nearest-neighbour imputation (impute::impute.knn, k = 10), and each retained protein was standardized to z scores before association testing. The final primary proteomic dataset included 37,698 participants and 2,915 proteins. Protein-level and participant-level missingness summaries, retention status and final analysis sample sizes are provided in Supplementary Data 3.

***Protein annotation for pathway and network analyses***

Olink assay identifiers were mapped to gene symbols for downstream annotation. Assays annotated to multiple genes were retained in PWAS, but their component gene symbols were used for enrichment analyses when a gene-set method required one symbol per gene. Ambiguous or non-gene identifiers were excluded from STRING network construction and recorded in the Supplementary Data.

***Functional enrichment analysis***

For pathway display, FDR-significant terms were ranked by adjusted P value within each gene-set collection. Representative terms were retained iteratively after excluding terms with high overlap with terms already selected. Gene-set overlap was measured by the Jaccard index, with a cutoff of 0.5. Up to ten representative terms were retained per collection for visualization; full enrichment results are provided in the Supplementary Data.

***STRING network analysis***

For STRING analysis^4^, only unambiguous gene symbols from Bonferroni-significant PWAS proteins were used. Assays annotated to multiple genes or non-gene identifiers were excluded from network construction and recorded separately. No additional interactors were added, so each retained node corresponded to a protein identified in the PWAS.

STRING edges were analysed as an undirected weighted network. Self-loops were removed, and duplicated protein pairs were collapsed by retaining the highest interaction score. In addition to maximal-clique centrality, node degree, weighted degree, betweenness centrality and k-core coreness were calculated for descriptive annotation. Louvain community detection was used to assign network modules.

***Protein age-trajectory analysis***

LOESS curves were fitted for each Bonferroni-significant PWAS protein across chronological age. Curves were evaluated on 61 equally spaced age-grid points across the analysed age range, using span 0.75 and polynomial degree 2. Proteins with insufficient non-missing observations or no measurable variation were excluded from trajectory fitting.

Protein trajectories were clustered using normalized protein-specific LOESS curves. Derived trajectory features captured early, middle and late age levels, early and late slopes, curvature, and the position of the maximum or minimum fitted value. These features were used for clustering and were not reported as association tests.

***DE-SWAN age-window analysis***

In the DE-SWAN analysis^5^, age was rounded to integer years and a six-year window was centred on each tested age. Participants in the younger half of the window were compared with participants in the older half of the same window, corresponding to [c - 3, c) versus [c, c + 3). Models were adjusted for sex, the first 20 genetic principal components, Townsend deprivation index, smoking status, alcohol intake frequency and BMI. For each age window, P values were adjusted across tested proteins using the Benjamini-Hochberg procedure. Proteins significant at retained crest ages were used for crest-specific enrichment analysis.

***MetScore2 Genome-wide association study***

Among participants in the primary MetScore2 cohort, 398,364 had available whole-genome sequencing data after genetic quality control. For the no-proteomics GWAS used in downstream proteome-wide MR and colocalization analyses, participants with baseline UKB-PPP Olink proteomic measurements were excluded before GWAS input generation. The final GWAS had a median variant-level sample size of 355,628 participants after variant-level genotype missingness and complete-covariate filtering.

REGENIE^6^ step 1 used a pruned set of genotype variants to account for relatedness and polygenic background. REGENIE step 2 tested chromosome-specific autosomal variants that passed genotype quality control under an additive model. Covariates were age, sex, age squared, age-by-sex interaction, age-squared-by-sex interaction, the first 20 genetic principal components and assessment centre. Sex and assessment centre were treated as categorical covariates. The median variant-level sample size was reported because sample size varied across variants after genotype filtering.

***LDSC clumping and fine-mapping***

For post-GWAS analyses, LDSC^7^ was restricted to HapMap3-overlapping variants. Independent lead variants^8^ were identified for reporting, whereas SuSiE RSS^9^ was used for fine-mapping within merged genome-wide significant loci. Locus-specific linkage-disequilibrium matrices were calculated from the same no-proteomics UK Biobank genotype subset used for the GWAS.

***Proteome-wide MR***

Protein-increasing alleles were aligned before MR analysis so that effect estimates represented the association of genetically higher plasma protein abundance with MetScore2. Variants were harmonized between the protein pQTL summary statistics and the no-proteomics MetScore2 GWAS by effect allele and non-effect allele. Ambiguous palindromic variants were removed when strand alignment could not be resolved.

The genome-wide significant cis-pQTL tier was used as the primary MR analysis. Cis-pQTL instruments were LD-clumped at r2 < 0.001 within a 10-Mb window, and instruments with F statistic ≤ 10 were excluded. Relaxed cis-pQTL tiers were used to assess consistency when additional instruments were allowed. The Wald ratio was used for single-instrument proteins, inverse-variance weighting for proteins with at least two instruments, and MR-Egger^10^, weighted median^11^ and weighted mode^12^ were used only when at least three instruments were available. Heterogeneity and pleiotropy statistics were treated as sensitivity checks.

***Colocalization analysis***

For colocalization, protein pQTL and MetScore2 GWAS variants were extracted from the same cis region and matched by GRCh38 chromosome, base-pair position and alleles. Colocalization used coloc.abf^13^ with priors p1 = 1 x 10^-4^, p2 = 1 x 10^-4^ and p12 = 1 x 10^-5^. Posterior probabilities were reported for the standard coloc hypotheses, with PP.H4 used to assess evidence for a shared causal variant.

***FinnGen disease Phe-MR***

The disease Phe-MR endpoint panel was curated from FinnGen R10 endpoints to cover age-related disease categories, including cardiovascular, cerebrovascular, metabolic, liver, renal, neurodegenerative, inflammatory, respiratory, musculoskeletal and cancer endpoints^14^. When several FinnGen endpoints represented closely related diagnoses, the endpoint with clearer clinical interpretation and adequate case count was prioritized. Endpoint names, FinnGen identifiers, case counts and control counts are provided in the Supplementary Data.

For each prioritized protein target, MR estimates were harmonized to the protein-increasing allele. Negative estimates for MetScore2 were interpreted as genetically higher protein abundance being associated with lower MetScore2, whereas positive estimates were interpreted as genetically higher protein abundance being associated with higher MetScore2. This direction was used to label the target-disease follow-up and was not treated as evidence that the protein can be safely modified therapeutically.

**Supplementary Data legends**

Supplementary Data 1. MetScore2 construction and model diagnostics, including mortality time-dependent AUCs, cohort flow, cross-fitted fold summaries, selected feature frequencies and SHAP feature summaries.

Supplementary Data 2. Biological-aging capacity analyses, including disease-onset comparisons, disease-risk prediction, MetScore1 comparison, frailty analyses, longitudinal MetScore2 change and modifiable risk-factor associations.

Supplementary Data 3. Proteome-wide association, enrichment and sex-sensitivity analyses, including PWAS results, enrichment results and sex-stratified PWAS outputs.

Supplementary Data 4. Proteomic interaction network outputs, including STRING network nodes, edges, hub rankings and module summaries.

Supplementary Data 5. Protein trajectory and DE-SWAN analyses, including trajectory clusters, cluster summaries, DE-SWAN crest summaries, crest proteins and crest enrichment results.

Supplementary Data 6. MetScore2 genome-wide association and fine-mapping outputs, including GWAS quality-control summaries, LDSC results, independent lead variants, locus summaries and SuSiE fine-mapping results.

Supplementary Data 7. Proteome-wide Mendelian randomization and colocalization outputs, including MR estimates, instrument summaries, colocalization results and prioritized protein evidence.

Supplementary Data 8. Target-disease Phe-MR analyses in FinnGen, including the endpoint panel, target instruments, full Phe-MR results, FDR-significant associations and quality-control summaries.

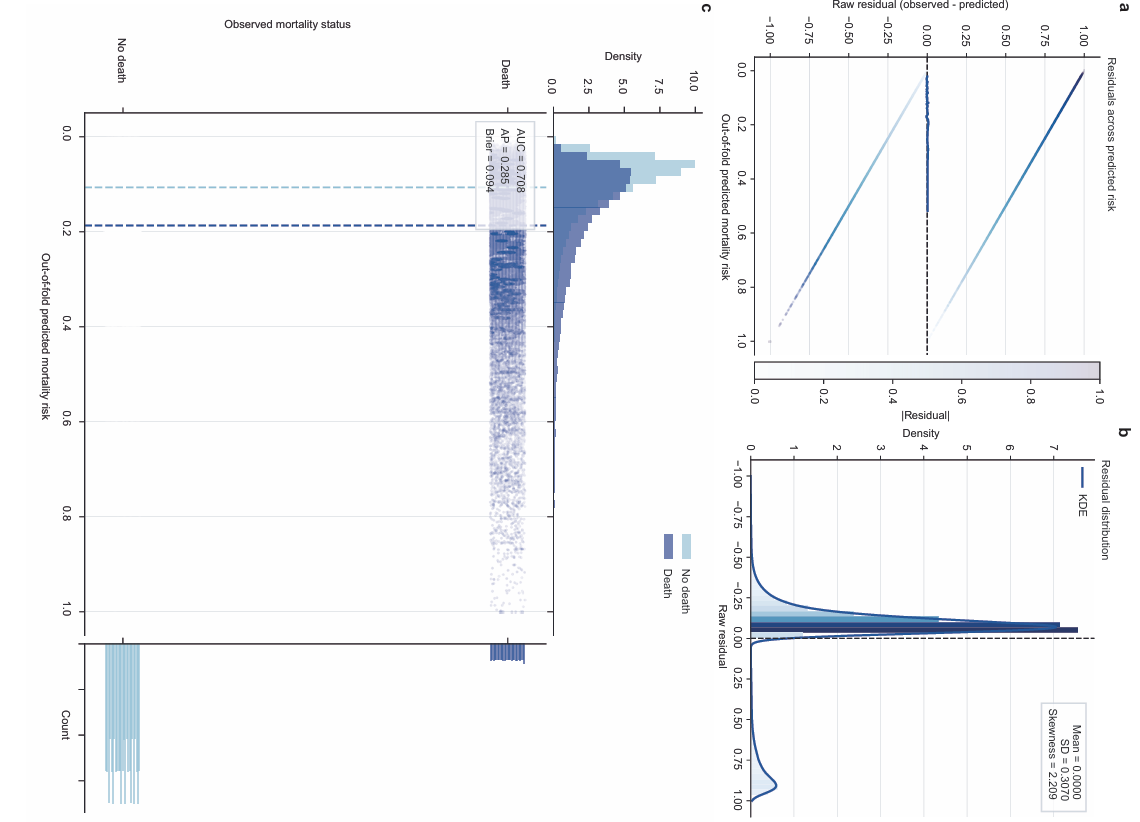
 **Supplementary Fig. 1 | Out-of-fold prediction and residual diagnostics for MetScore2.**

**a,** Distribution of out-of-fold linear predictors across participants, stratified by mortality status. **b,** Distribution of prediction residuals, used to assess whether residual structure remained after model fitting. **c,** Mean observed mortality and residual summaries across quantiles of the out-of-fold score. Points show bin-level estimates and intervals show the corresponding uncertainty where plotted. The diagnostics were generated from out-of-fold predictions so that each participant was scored by a model that was not trained on that participant.

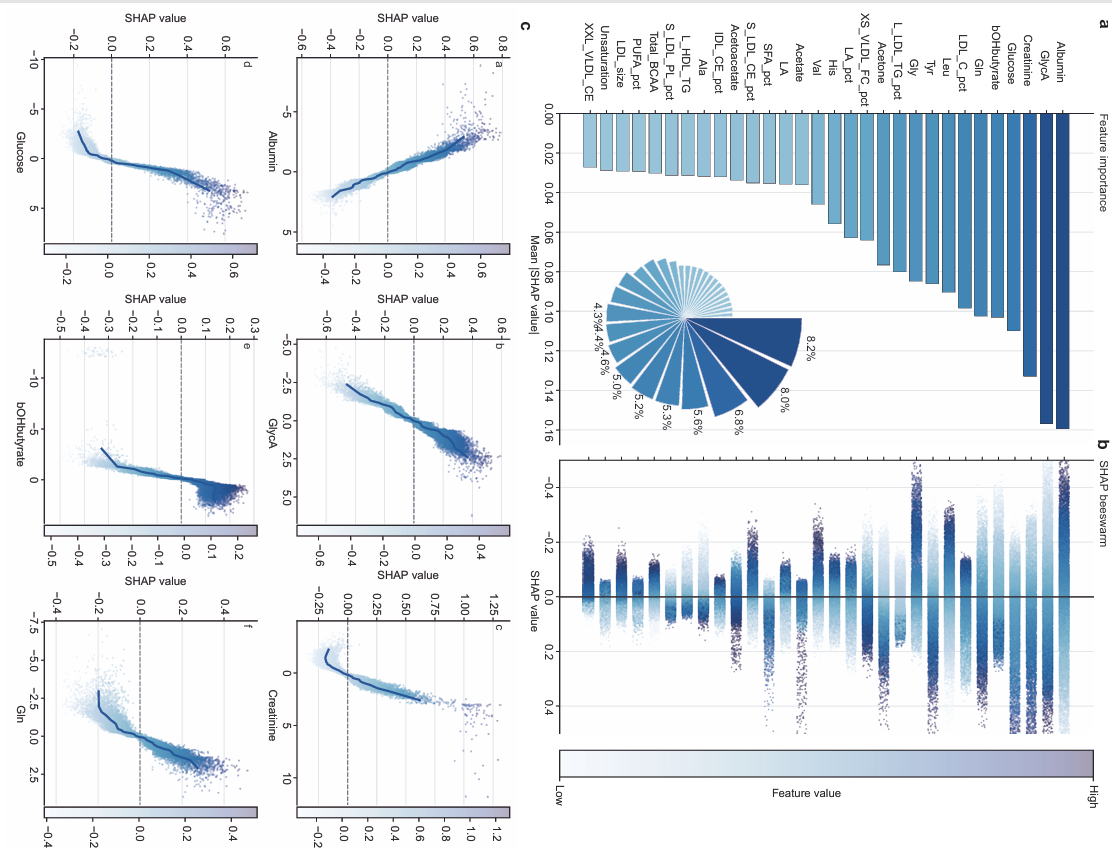
 **Supplementary Fig. 2 | Metabolite contributions to MetScore2 predictions.**

**a,** Mean absolute SHAP values for the leading NMR metabolic features used by MetScore2. The inset pie chart shows each displayed feature’s share of total importance among the plotted metabolites. **b,** SHAP beeswarm plot showing the direction and distribution of feature contributions across participants. Positive SHAP values increase the predicted mortality risk, whereas negative values decrease it. Point colour denotes the standardized metabolite value. **c,** SHAP dependence plots for albumin, GlycA, creatinine, glucose, beta-hydroxybutyrate and glutamine. Points represent participants, curves show fitted dependence trends, and dashed horizontal lines indicate zero contribution.

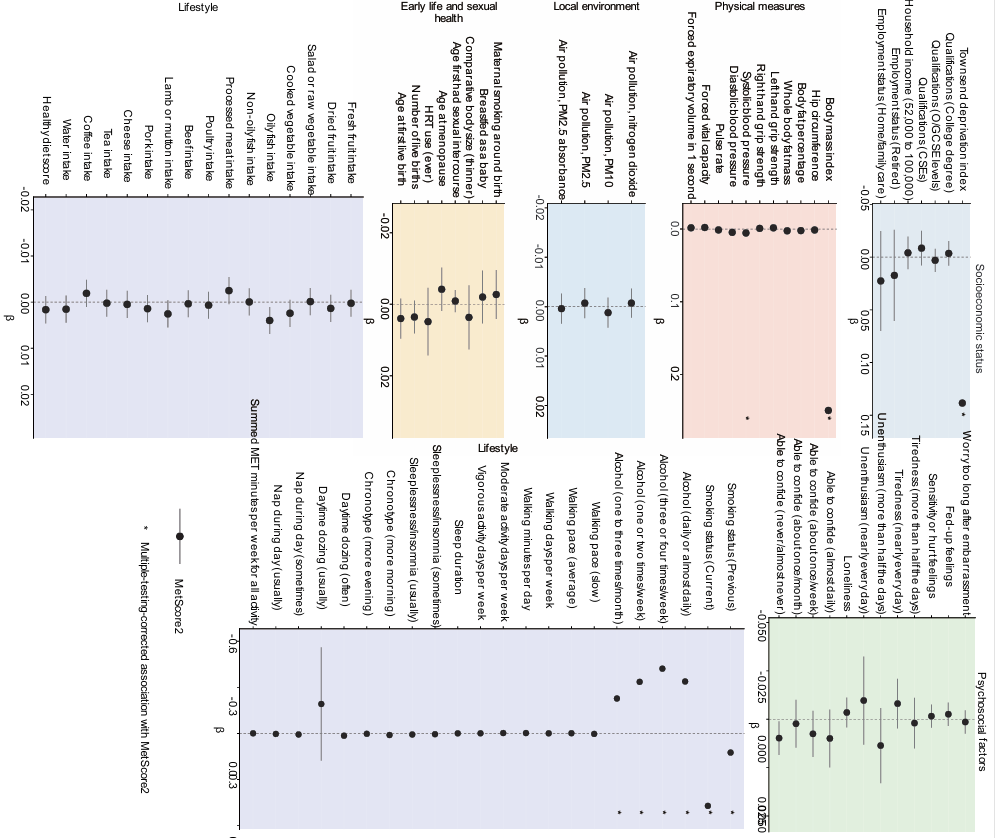
 **Supplementary Fig. 3 | Associations between MetScore2 and modifiable clinical, social and lifestyle factors.**

Regression estimates for selected potentially modifiable correlates of MetScore2, grouped by domain. Effect estimates are shown as beta coefficients with 95% confidence intervals after adjustment for the covariates specified in the Methods. Domains include socioeconomic status, physical measures, local environment, early-life and sexual-health variables, diet, psychosocial factors and lifestyle behaviours. Positive estimates indicate higher MetScore2 among participants with higher levels of the exposure or the specified exposure category. Multiple-testing-adjusted significance is indicated where shown. Full estimates and variable definitions are provided in the source data.

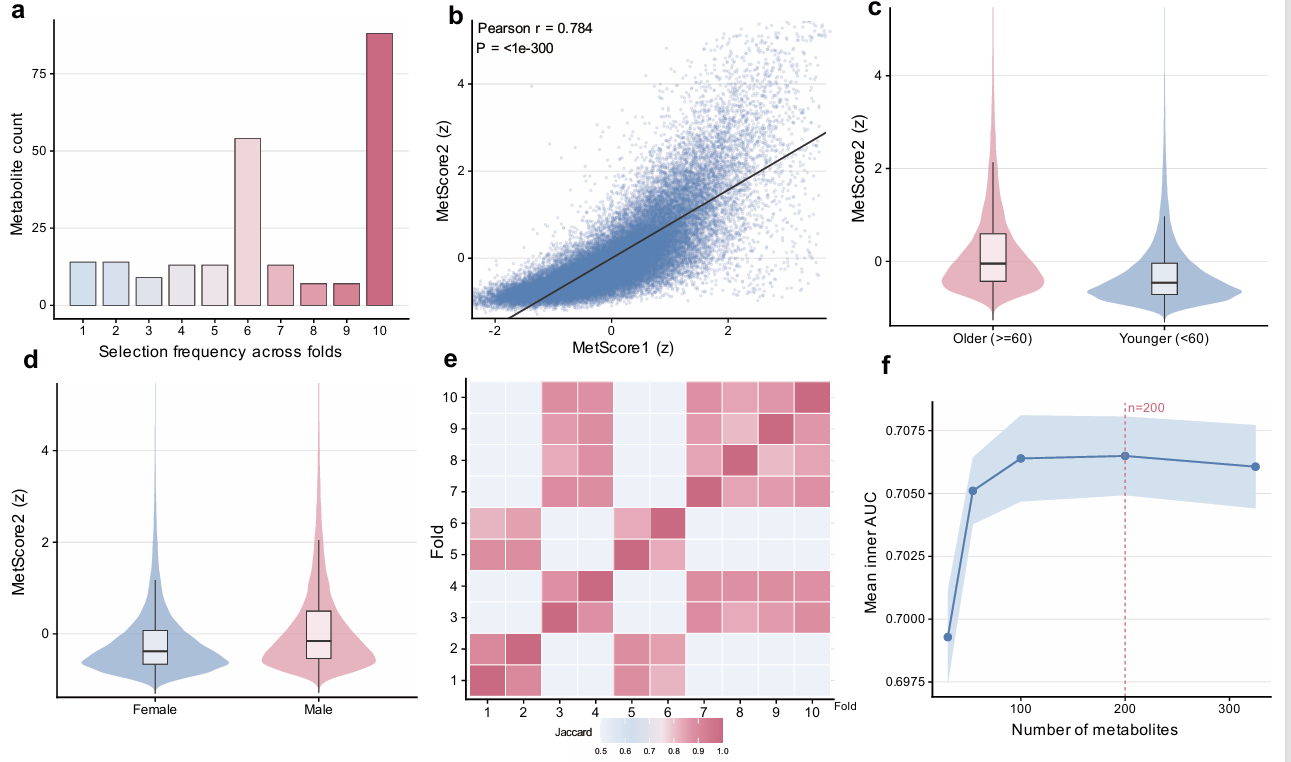
 **Supplementary Fig. 4 | MetScore2 feature-selection stability and score distributions.**

**a,** Selection frequency of metabolites across the ten outer cross-validation models. **b,** Agreement between MetScore2 and the earlier MetScore1 in participants with both scores available. **c,d,** Distributions of MetScore2 by age group and sex. **e,** Pairwise Jaccard similarity of selected metabolite sets across outer folds. **f,** Inner-loop model performance across candidate numbers of selected metabolites; The selected feature number was 100 in four outer folds and 200 in six outer folds. Correlation in b is Pearson's r. Box plots show the median, interquartile range and whiskers defined by the plotted data. Full feature-selection results are provided in Supplementary Data 1.

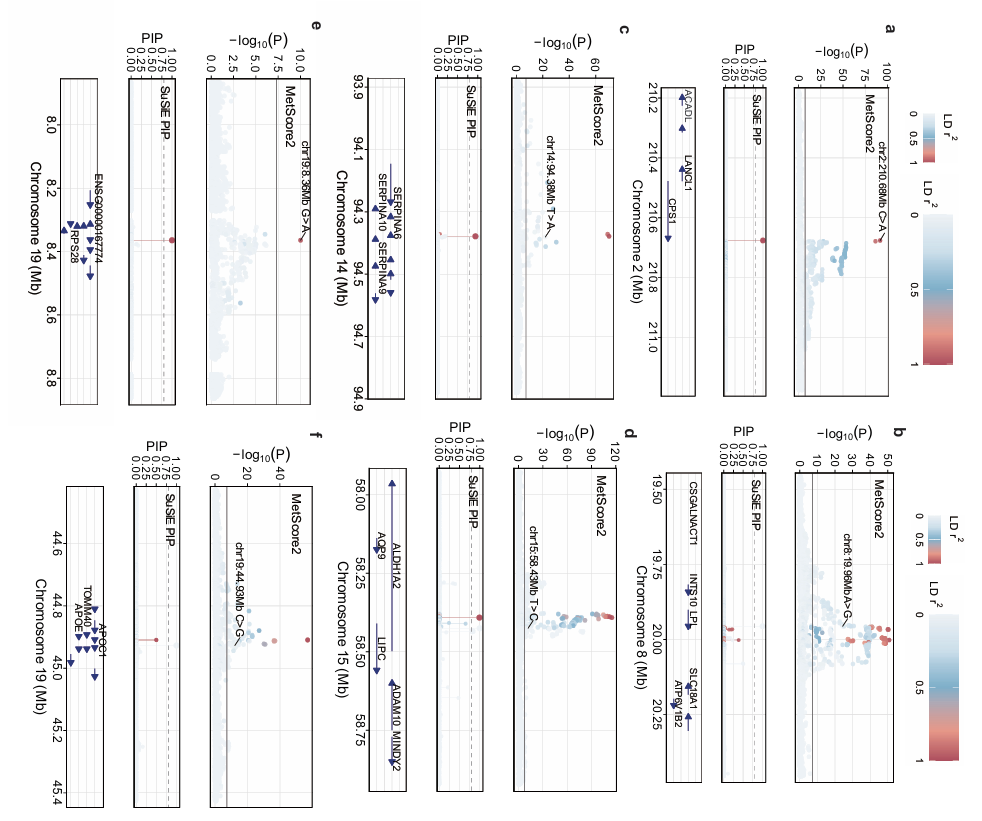
 **Supplementary Fig. 5 | Regional fine-mapping of representative MetScore2 GWAS loci.**

Representative genome-wide association and fine-mapping results for MetScore2. **a-f,** Regional association plots at selected loci, with variants coloured by linkage disequilibrium to the lead variant where shown. Panels include SuSiE credible-set summaries and posterior inclusion probabilities for variants with the strongest fine-mapping support. Association testing used WGS variants and the MetScore2 phenotype under the genetic analysis model described in the Methods. The displayed loci illustrate distinct regional architectures among the 78 genome-wide significant loci identified for MetScore2. Complete GWAS, lead variant and fine-mapping results are provided in Supplementary Data 6.

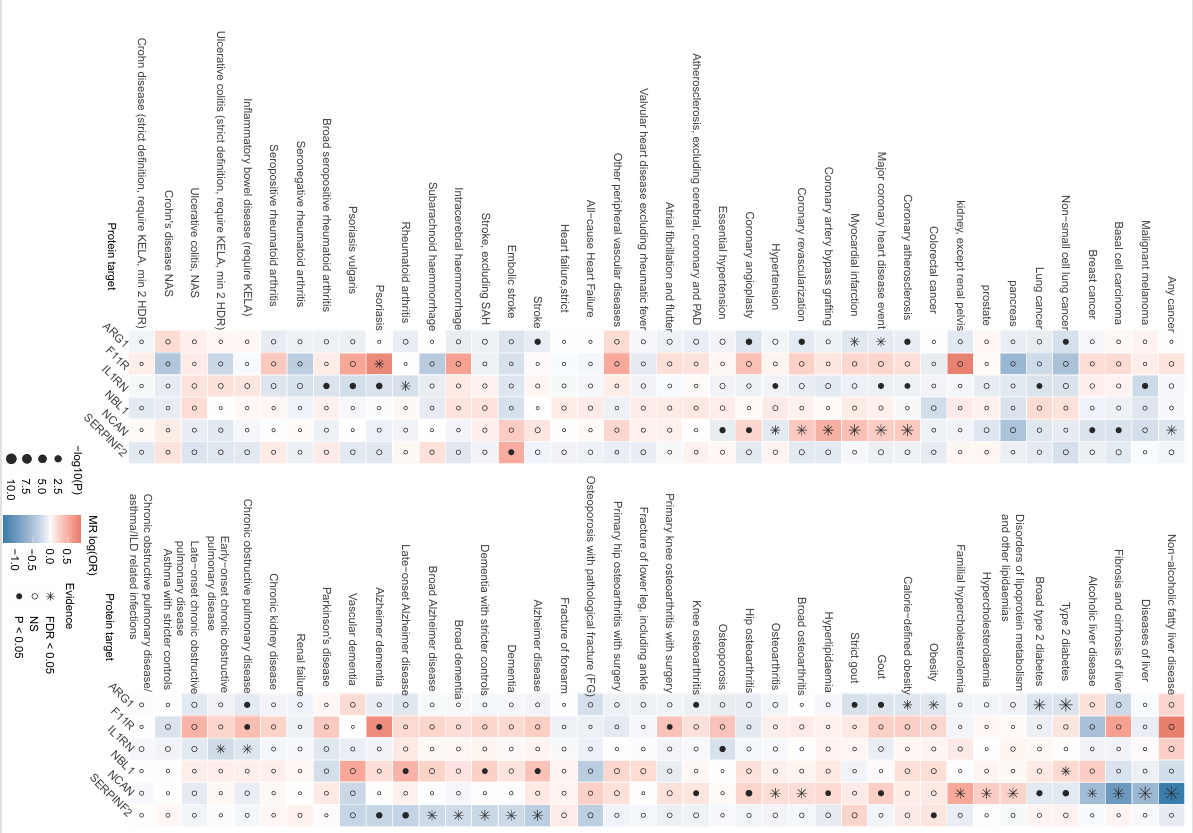

**Supplementary Fig. 6 | FinnGen phenome-wide Mendelian randomization profile of prioritized targets.**

Heat map showing Mendelian randomization associations between six prioritized protein targets and 80 age-related disease endpoints in FinnGen. Rows represent protein targets and columns represent disease endpoints grouped by clinical domain. Colour denotes the direction and magnitude of the log odds-ratio estimate where shown, and point size or symbols denote statistical support according to the plotted scale. The analysis evaluated 480 target-disease pairs and identified associations after global false-discovery-rate control. Complete endpoint definitions, effect estimates, confidence intervals, P values and FDR-adjusted results are provided in Supplementary Data 8.
